# Multi-omic Evaluation of Metabolic Alterations in Multiple Sclerosis Identifies Shifts in Aromatic Amino Acid Metabolism

**DOI:** 10.1101/2020.12.30.20249031

**Authors:** Kathryn C. Fitzgerald, Matthew D. Smith, Elias S. Sotirchos, Michael D. Kornberg, Morgan Douglas, Bardia Nourbakhsh, Jennifer Graves, Ramandeep Rattan, Laila Poisson, Mirela Cerghet, Ellen M. Mowry, Emmanuelle Waubant, Shailendra Giri, Peter A. Calabresi, Pavan Bhargava

## Abstract

The circulating metabolome is a product of interactions between the genome, epigenome, exposome and microbiome. The metabolome may be altered in people with multiple sclerosis (MS); however, existing metabolomics studies were relatively small or characterized a limited number of metabolites. Herein, we performed a multi-site study profiling the circulating metabolome to obtain relative abundances for 269 metabolites in a large cohort of MS patients and healthy controls. After adjusting for batch effects and extensive quality control, we created an overall metabolic dysfunction score, defined apriori sets of metabolites using known metabolic pathways, and derived novel networks of correlated metabolites using a weighted correlation network analysis (WGCNA). We assessed whether metabolic dysfunction, individual metabolites, metabolic pathways or WGCNA-identified module scores differed between people with MS versus healthy controls (HC) after adjusting for age, sex and race using generalized estimating equations (participants could provide multiple samples). In a subset of patients, information on disability status was also available. Similar models assessed the association between metabolites and metabolite sets with measures of disability. In people with MS, we identified striking abnormalities in a WGCNA-defined module enriched in aromatic amino acid (AAA) metabolites (FDR-adjusted p-value=2.77E-18) that are also strongly associated with disability (FDR-adjusted p-value for AAA module=1.01E-4). Consistent results were obtained using apriori-defined metabolite sets or in analyses of individual metabolites. The identified abnormalities likely relate to imbalances in gut microbial metabolism of AAAs resulting in reduced production of immunomodulatory metabolites and increased production of metabotoxins (indole acetate, phenylacetylglutamine, p-cresol sulfate, p-cresol glucuronide). Single cell RNA sequencing data analysis demonstrated altered AAA metabolism in CSF and blood derived monocyte cell populations, while treatment of human peripheral blood mononuclear cells with AAA-derived metabotoxins resulted in increased production of tumor necrosis factor-α. We identify novel metabolic alterations in people with MS potentially contributing to disease pathophysiology.

## INTRODUCTION

Multiple sclerosis (MS) is an inflammatory and neurodegenerative disorder of the central nervous system (CNS) (1) The etiology of MS is multifactorial and likely involves multiple levels of biological interactions with genetic and environmental contributors (2, 3). However, detailed understanding of these interactions or underlying mechanisms remain rudimentary. Molecular profiling of circulating small molecules using metabolomics integrates many of these systems, as an individual’s metabolic phenotype reflects an intersection between environmental sources of variation such as lifestyle characteristics (e.g. diet), upstream genetic influences and activity of the gut microbiota (4, 5). Thus, the assembly of a detailed global map of functional relationships in MS using circulating biologic intermediates may provide a valuable step forward to improve our understanding of potential contributors to MS pathogenesis.

Preliminary studies suggest that metabolic alterations exist in people with MS with respect to global metabolomic profiles, specific pathways and individual metabolites (6–9). These results are consistent with observations from metabolomics studies of other neurological disorders, including Alzheimer’s and Parkinson’s disease, and other autoimmune disorders (10–12). However, with respect to MS, these studies have been restricted to small patient populations and limited arrays of metabolites. And critically, prior studies have not evaluated the links between metabolomic differences and patient characteristics, including measures of disease severity or other candidate biological drivers of disease.

In the current study, we compare nearly 1000 detailed metabolomic profiles from people with MS and healthy people from across the age spectrum using individual metabolite and pathway-level analyses. Our findings highlight distinct abnormalities in aromatic amino acid metabolism that imply an altered balance of immunomodulatory metabolites (e.g. arylhydrocarbon receptor [AhR] and hydroxycarboxylic acid-3 [HCA_3_] receptor agonists) and a shift toward production of known metabotoxins (e.g. indole acetate, phenylacetylglutamine, p-cresol sulfate/glucuronide). Metabolomic alterations, particularly in these immunomodulatory amino acids and metabotoxins, were associated with disease severity. We then integrated single-cell transcriptomics data to identify alterations in genes involved in aromatic amino acid metabolism in MS peripheral blood and CSF derived immune cells. Lastly, we tested the effects of the identified metabotoxins on human peripheral immune cells and noted changes in the production of the pro-inflammatory cytokine TNF-α from monocytes, suggesting functional roles for these metabolites that are altered in MS and are related to disease severity.

## METHODS

### A. Study Population

Study participants were pooled from three sites: Johns Hopkins MS Center (JHU), the University of California at San Francisco (UCSF), and the Henry Ford Hospital MS Center or Accelerated Cure Project (ACP; samples were acquired by Henry Ford).

#### A.1 JHU

JHU study participants (n=640) were pooled from several ongoing or recently completed clinical research studies in which blood samples were acquired for metabolomics analyses. We included baseline metabolomics samples (e.g. before the initiation of the intervention) from two recently completed studies of dietary interventions in people with relapsing-remitting MS: one study included relapsing-remitting MS patients aged 18-75, with BMI ≥25 kg/m^2^ and receiving natalizumab,(13) the other included relapsing-remitting MS patients, aged 18-50, untreated or treated with injectable MS therapies (interferon beta or glatiramer acetate) and a BMI ≥22.5 kg/m^2^ (13, 14). We also included samples from an observational study of relapsing-remitting MS patients who were treated with dimethyl fumarate and age- and race-matched healthy controls (HC;) (15). Finally, we included baseline samples from a study of Caucasian age-matched relapsing-remitting MS patients and HC who had serum 25-hydroxyvitamin D levels <20ng/mL (16). For remaining studies, participants with MS (including both relapsing-remitting and progressive subtypes) and HC were recruited by convenience sampling from the JHU MS Center and provided blood samples. Blood was processed within 3 hours of collection using a standardized protocol that was common to all the studies conducted at JHU, and serum or plasma was aliquoted and stored at - 80 C° until metabolomics analyses.

##### A.1.1 Disease severity measures

In a subset of study participants (n=305), information on measures of disease severity, including measures of disability status (via the Expanded Disability Status Scale [EDSS]), were available. A subset of participants (n=192) also had optical coherence tomography (OCT) scans, as an imaging biomarker of retinal neurodegeneration that were obtained within 6 months of metabolomics assessment. Segmentation of retinal layer thicknesses was performed using a validated automated segmentation algorithm, as previously described.(17–19) We assessed the association between metabolite levels and thickness of the macular ganglion cell + inner plexiform layers (GCIPL); we selected GCIPL thickness as the primary OCT phenotype as this composite measure is more strongly associated with brain atrophy and is less vulnerable to swelling that may occur in the context of inflammation of the optic nerve (18, 20). Eligible participants were those who did not have a history of diabetes mellitus, uncontrolled hypertension, glaucoma, prior ocular surgery or trauma, refractive errors exceeding ±6 diopters, or other significant neurological or ophthalmological conditions.

#### A.2 Henry Ford / ACP Samples

Individuals with MS (including both relapsing-remitting and progressive subtypes) as well as HC were recruited by convenience sampling from the Henry Ford Hospital MS Center. A subset of MS serum samples was acquired from serum repository at the Accelerated Cure Project (ACP). For both sets of MS samples, participants (n=184 total) had to have a confirmed diagnosis of MS and provide blood samples, which were used for metabolomics assessment. Metabolomics assessment was conducted in a single batch, regardless of the original parent study (e.g. Henry Ford or ACP). Blood samples from HFH MS Center and ACP were obtained through an IRB approved study of the metabolomics signature in MS.

#### A.3 UCSF Pediatric Cohort

MS patients were selected randomly from two cohort studies of pediatric MS led by investigators at UCSF (n=136). These cohorts included: 1) pediatric MS or clinically isolated (CIS) patients enrolled in a prospective cohort study from the UCSF Pediatric MS clinic, 2) pediatric MS or CIS patients who participated in a multi-center case-control study evaluating risk factors for pediatric MS and who also provided demographic and clinical information (9, 21, 22). For both cohorts, eligible MS participants were those with relapsing-remitting MS or CIS with high risk of conversion to MS (e.g. ≥ 2 T2 hyperintense foci on MRI), with disease onset <18 years and who had seen a neurologist within 4 years of symptom onset (23). Diagnoses were confirmed by a pediatric MS specialist. HC were recruited from general or specialty pediatric clinics and were aged <22 years, did not have a history of autoimmune disorders (except asthma or eczema) or previous history of severe health condition or treatment with immunosuppressive medications or have a biological parent or sibling with MS. All participants provided a blood sample at the time of recruitment into the study. Disability status via the EDSS score was also available on people with MS. A subset of participants also provided a stool sample. Blood was processed and frozen at −80 C° within 3 hours of collection.

### B. Assessment of metabolomic profiles

All metabolomics analyses via mass spectrometry were conducted at Metabolon (Durham, NC), and methods have been described in detail elsewhere (7). JHU samples were pooled from metabolomics analyses conducted in 7 different batches, while UCSF and Henry Ford samples were each analyzed in separate single batches, totaling 9 batches overall. Briefly, samples were thawed and underwent additional preparation (derivatization), as previously described. The derivatized samples were subjected to either gas chromatography followed by mass spectrometry (GC/MS) or liquid chromatography followed by tandem mass spectrometry (LC/MS/MS). Mass spectra obtained from these techniques were then matched to a library of spectra derived from standards to identify specific metabolites, and the area under the curve for the mass spectra was used to calculate the relative abundance of each metabolite.

### C. Single cell analysis of blood and cerebrospinal fluid (CSF) in MS vs. HC

In follow-up analyses, we also assessed whether any metabolic differences we observed in the serum or plasma correlated with changes in gene expression at the cellular level. To do so, we used publicly available single cell RNA sequencing (snRNA-seq) data from an existing study comparing cellular-level differences in metabolic gene expression from an integrative analysis of blood and CSF from MS patients and HC (24). Eligible MS patients for the original study were treatment-naïve patients with an initial episode suggestive of MS (CIS) or those with RRMS. Eligible control subjects were those providing blood and CSF samples following workup for idiopathic intracranial hypertension (IIH) matched to MS subjects by age, sex, and CSF features (protein, lactate, and glucose levels); full details are provided in Schafflick et al.(24)

### D. Functional effects of AAA-derived metabotoxins on human monocytes

To determine whether AAA-derived metabotoxins affect human immune cell function, we isolated peripheral blood mononuclear cells (PBMCs) from healthy controls and cultured them for 24 hours with either vehicle or varying doses of indole acetic acid (IAA) or phenylacetylglutamine (PAG). At the end of this time period we stained cells with surface antibodies followed by washing and then we permeabilized cells and stained for intracellular markers. Cells were then acquired on an Aurora Cytek flow cytometer. We analyzed effects on CD14+ monocytes – monocyte subsets (classic, intermediate and non-classical), activation markers (CD83, CD86), and cytokine production (IL6, TNF-a). We then analyzed the effects of these metabolites on human monocytes using a one-way ANOVA to compared markers between the different treatment groups

### E. Statistical Analysis

#### E.1. Quality Control

We initially included 329 metabolites that were measured in each batch across 9 metabolomics runs in 960 total samples. We then implemented the following quality control (QC) procedure to identify potential outlying metabolites and/or samples. First, we removed metabolites with >20% missing values across samples (n=27) and imputed missing metabolite values using k-nearest neighbors (10 neighbors used for each imputation); consistent results were observed when using the minimum value of observed metabolites. We then log-transformed all metabolites. We adjusted for batch (and site/specimen type) using the ComBat algorithm, which is a harmonization technique originally developed for the analysis of microarray data and was designed to remove batch-related extraneous variation while conserving biological variation (25, 26). Results before and after the application of ComBat are displayed in **Supplemental Figure 1**. Our metabolomics analyses also included both between-batch replicates (e.g. the same sample collected on the same day was included in multiple metabolomics runs; n=80 samples) and within-batch replicates (e.g. the same sample was included twice in the same metabolomics run; n=70 samples). We calculated the intraclass correlation coefficient (ICC) for both between- and within-batch replicate samples. ICCs for both within- and between-batch were generally high; the median ICC for within-batch replicates was 0.94 (IQR: 0.86, 0.98) and the median for between-batch replicates was 0.79 (IQR: 0.63, 0.89; **Supplemental Figure 1**). We removed metabolites which had ICCs < 0.40 in 1) between batch replicates (n=27 metabolites), 2) within-batch replicates (n=4 metabolites) or 3) both (n=2 metabolites). The final analyses included 269 metabolites. From metabolomic samples with within- or between-batch replicates, we randomly selected one from each set to be eligible for inclusion in the main analyses. We tested for potential sample outliers using principal components analyses (PCA); we excluded those who were > 3 standard deviations (SD) of the mean for PC1 and PC2 (n=1 excluded). We also identified potential outliers using a Euclidean distance-based sample network (and a standardized network connectivity measure [Z_k_], as described in Horvath et al.); outlying samples were classified as those who had Z_k_ values < −4 (n=5 samples), as suggested by Horvath et al; leaving 954 samples (99.1%; n=756 unique individuals) eligible for the analysis (27).

We also compared the distribution of within-person and between-person dissimilarity in overall metabolomic profiles, as several participants in our study contributed multiple samples. We calculated the within-person dissimilarity as the median Mahalanobis dissimilarity between metabolomic profiles taken from the same individual (28). To calculate between-person dissimilarity, we first averaged metabolomic profiles taken from the same individual to estimate a composite “average” metabolomic profile. We then calculated the median Mahalanobis dissimilarity between metabolomic profiles from different individuals.

#### E.2. Main analyses comparing metabolomic profiles between MS patients and HC

For our primary analyses, we compared metabolomic profiles between people with MS and HC considering 1) global differences in the overall metabolome, 2) differences in individual metabolites, and 3) differences in composite metabolic pathway measures.

##### E.2.1. Metabolomic dysfunction score

To identify samples with highly divergent metabolomic composition, we created an age-adjusted global “metabolomic dysfunction” score based on Mahalanobis dissimilarities between MS patients and HC. To do so, we set the ‘reference population’ as all age- and sex-adjusted metabolomic profiles from HC and derived a metabolomic dysfunction score as the median Mahalanobis dissimilarity among age- and sex-adjusted metabolomic profiles in MS patients to this reference set. To identify samples which were highly divergent from the reference population, we thresholded the metabolomic dysfunction score at the 90^th^ percentile (e.g. samples which have metabolomic feature configurations that have <10% probability of occurring in a person without MS; (28). Sensitivity analyses also adjusted for race/ethnicity in metabolomic dysfunction scores.

##### E.2.2. Individual metabolite and metabolic pathway-based analyses

We compared individual log-transformed metabolites between people with MS and HC using generalized estimating equations (GEE; to account for multiple metabolomic profiles contributed by some participants) with a Gaussian link function and adjusted for age, sex, and race/ethnicity. Additional analyses adjusted for body mass index (BMI; kg/m^2^), which was only available for a subset of individuals (n=568). For metabolic pathway-based analyses, we performed two sets of complementary analyses: an agnostic approach to discover sets of related metabolites and another incorporating known information on metabolic pathways and metabolite interactions. For the agnostic approach, we derived novel sets of related metabolites using a weighted gene-expression correlation network analysis (WGCNA);(27) WGCNA is a systems biology-based technique that was originally developed to study correlation patterns in gene expression and has been extended to other settings (including our previous work in metabolomics), cancer, and neuroimaging (7, 15, 29). The goal of WGCNA is to find clusters (or metabolic modules in this case) of highly interconnected nodes (metabolites, here) using a correlation network. The identified metabolic modules can be summarized into single measures (i.e. eigen-metabolites) as the first principal component of the identified module and can be used in subsequent analyses. For a priori-based pathway analyses, we performed two sets of related analyses incorporating known biological information. For the first, we classified metabolites into groups (>3 metabolites) based on related biologic function (e.g. glutathione metabolism, tryptophan metabolism, among others); pathway membership for each metabolite are included as a part of **Supplemental Tables 2, 3 and 7**. We applied a resampling-based permutation-based algorithm to assign statistical significance while preserving metabolite-metabolite correlation. To do so, we fit individual models for each metabolite (via GEE, adjusting for age, sex and race/ethnicity, as above) and extracted and ranked the obtained p-value; ranks for a given pathway set were then averaged. We then permuted phenotype labels 10,000 times and repeated the above procedure to calculate an average rank of the p-values for a pathway set for each permutation. Finally, the p-value for a given pathway set is the probability that the observed average rank is less than the expected average rank (as calculated from the 10,000 permutations). For the second, we used known metabolic reactions and interactions from available databases to create a metabolic network to use for subsequent analyses. To do so, we downloaded all MetaCyc compounds and corresponding reactions (30). Individual compounds served as the nodes, and we set an edge to be a known (or predicted) reactant-product metabolic interaction; in total, the network included 2248 nodes (metabolites) and 7449 edges (reactant-product metabolic interaction). We then mapped results of the individual metabolites that are potentially different between MS patients and HC (e.g. individual metabolites which differed between MS vs. HC with FDR <0.25) and extracted subnetworks of enriched metabolite interactions (corresponding roughly to metabolic pathways) using Prize-collecting Steiner Forest (PCSF) graph optimization (31). We also performed analyses assessing pairwise ratios of measured metabolites on opposite sides of MetaCyc-defined metabolic reactions, as a proxy for estimated enzyme activity of metabolic reactions. For this analysis, we included 139 metabolite-metabolite ratios participating in 207 metabolic reactions and tested for differences between MS and HC using multivariable-adjusted GEE, as above.

##### E.2.3. Sensitivity analyses for metabolite and metabolite pathway analyses

We performed an additional set of sensitivity analyses where repeated analyses using a leave-one-out procedure where we excluded individual batches and repeated all analyses in the remaining set to ensure that a single subgroup of samples was not driving the observed associations. Sensitivity analyses additionally stratified by serum and plasma origin of each individual sample and pooled results using meta-analysis methods.

#### E.3 Association between metabolomic profiles and disease severity measures

We assessed the association between EDSS (417 EDSS scores from 312 individuals with MS) and metabolomic profiles (i.e. global metabolomic differences, individual metabolites, and pathway-based analyses) and metabolite ratios using multivariable-adjusted GEE, as above. In sensitivity analyses, we also assessed the age-related MS severity score (ARMSS; as a marker of MS severity) and metabolomic profiles (and did not adjust for age in these analyses). ARMSS is similar to the MS severity score, except it uses age instead of disease duration, as age is typically unbiased and more readily obtained than disease duration. In the subset of individuals with OCT imaging linked to a metabolomics sample (n=192), we also assessed whether GCIPL thickness (as a quantitative imaging measure of retinal neurodegeneration) was associated with selected metabolites (or metabolite ratios) that demonstrated significant differences between MS and HC (metabolites with FDR-adjusted p<0.05 among the 269 individual metabolite tests). Similar to above, we used GEE (to account for multiple eyes per person) and adjusted for a similar set of covariates. Similar sensitivity analyses additionally stratified by serum and plasma origin of each individual sample and pooled results using meta-analysis methods.

#### E.4 Changes in metabolic gene expression in single cell analysis of blood and cerebrospinal fluid (CSF) in MS vs. HC using pathway activity scores

After downloading raw cell counts, we implemented a standard quality control procedure (e.g. excluding low quality samples with few detected cells and features, cells with fewer than 200 features [gene], cells with ≥2500 features, cells with <5% of percentage of all features coming from mitochondrial gene sets,). Normalized samples were integrated using the Seurat pipeline v3 (e.g. selection of integration anchors using canonical correlation analysis and 30 PCs). Samples were then clustered using a shared nearest neighbor (SNN) modularity optimization-based clustering algorithm; clustering and sample integration were then inspected visually for quality and separation. Based on marker gene expression, we identified a several clusters of T cells: effector-memory (EM)-like CD4+ T cells (*CD69*), central-memory (CM)-like CD4+ T cells (*CD27*), naïve CD4+ T cells (*TRAC, CD4*), activated CD8+ cells (*CD8B* and *CCL5*), non-activated CD8+ cells (*CD8B* and *CCR7*) and T-regulatory cells (*FOXP3*). We also identified clusters of NK cells (*GNLY, NKG7*), B-cells (*CD79A*), and plasmablasts (IGHG). We also identified monocyte clusters and monocyte cell clusters that were mostly blood-derived (Mono-B; *FCGR3A/CD16*) or CSF-derived (Mono-*CSF*; *CD14, C1QA, C1QB*). Other myeloid lineage cells were clustered into mDC type 1 (mDC1; WDFY4, BATF3), mDC type 2 (mDC2; *FCER1A, CD1C*). Other clusters included plasmacytoid dendritic cells (pDC; TNFRSF21) and megakaryocytes (*MegaK; GNG11*). We detected one cluster of red blood cells (*HBA1, HBA2*, and *HBB*), which was removed from the analysis. tSNE plots following sample clustering and labeling are provided as **Supplemental Figure 2**). Our primary goal for this analysis was to identify cell-type specific shifts in metabolic gene expression occurring between MS and HC at the pathway level. To do so, we calculated a cell-type specific pathway activity score using curated gene/pathways lists ways (including pathways with at least 10 genes) that are available in Xiao et al. and adapting a similar methodology described therein (32). Briefly, the pathway activity score is calculated through the following steps 1) calculate the mean expression level for gene *i* for cell type *j* for *k* individuals as 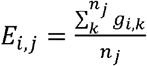 and the average gene expression across all cell types, 2) calculate average relative expression for a given cell type relative to the average expression across cell types as 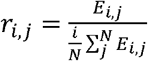, 3) derive final pathway activity score for *t* pathways as the weighted average of cell type-specific relative expression values, *r*_*i,j*_, for each participating gene as 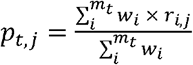. Weights are derived as the number of pathways in which a gene participates. Pathways activity scores for each cell type were averaged for MS and HC and stratified by sample type (blood, CSF) separately. In analyses, we compared the ratio between MS and HC 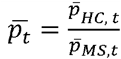. Statistical significance was assessed by a permutation test of sample labels.

## RESULTS

We included 954 metabolomic profiles from 756 individuals with MS or HC (514 MS patients and 214 HC) in which 269 metabolites were reliably measured (**Supplemental Figure 1**). Participants were aged 40.26 ± 15.46 years (**Table 1; Supplemental Table 1; Figure 1**) and were predominantly female (73.49%) and non-Hispanic whites (84.21%). With respect to MS, the majority of participants had relapsing-remitting MS (72.71%), disease duration was 13.45 ± 10.63 years, and were, on average, moderately disabled (median EDSS score = 3.0; 27% reported using a cane).

**Figure 1.**
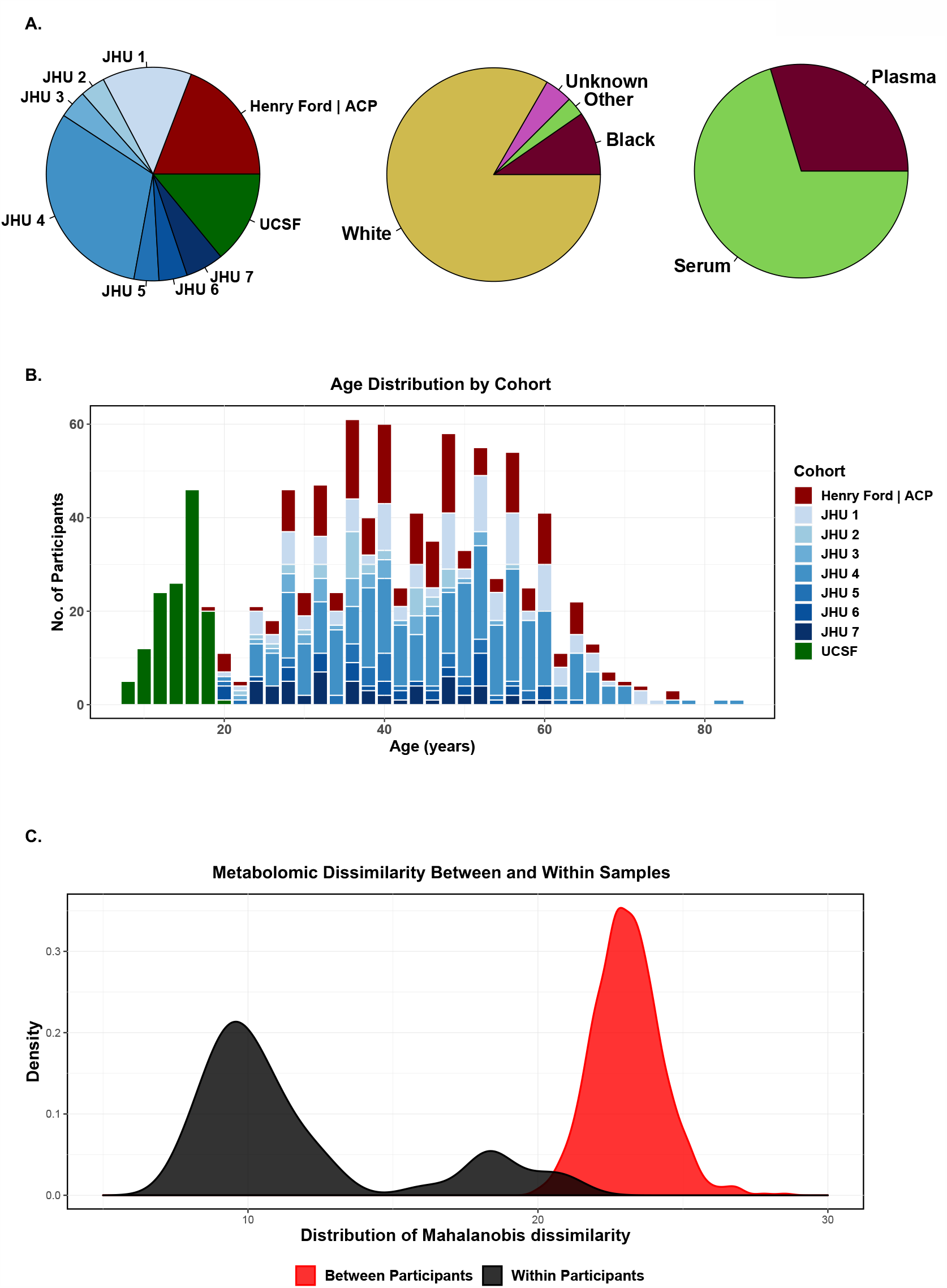
Demographic characteristics of included study participants. **A**. The proportion of participants samples by different sites, races and sample media type. **B**. Age distribution of included samples colored by contributing sites. **C**. Distribution of Mahalanobis dissimilarity for between and within-person samples. Within-person dissimilarity is calculated as the median Mahalanobis dissimilarity between metabolomic profiles taken from the same individual. Between-person dissimilarity is calculated as the median Mahalanobis dissimilarity between metabolomic profiles from different individuals. As expected, between-person dissimilarity was significantly greater than within dissimilarity (p<1E-16).

**Table 1.** Characteristics of included study participants.

| **Disease Status** | **HC** | **MS** |
| --- | --- | --- |
| **No. samples** | 317 | 637 |
| **N** | 241 | 515 |
| **Age, years, mean (SD)** | 35.85 (15.71) | 42.54 (14.91) |
| **Age range (min, max)** | 7, 84 | 7, 76 |
| **Female sex, n (%)** | 224 (70.66) | 468 (73.47) |
| **Caucasian race, n (%)** | 253 (79.81) | 542 (85.09) |
| **Disease duration, years, mean (SD)** |  | 13.45 (10.63) |
| **Progressive MS, n (%)** |  | 205 (32.18) |
| **EDSS (417 scores available), median (IQR)** |  | 2.50 (1.50, 5.00) |
| **Use of cane, n (%)** |  | 97 (23.2) |
| **Disease modifying therapy, n (%)** |  |  |
| no treatment |  | 195 (30.61) |
| interferon beta |  | 60 (9.42) |
| glatiramer acetate |  | 130 (20.41) |
| dimethyl fumarate |  | 36 (5.65) |
| teriflunomide |  | 2 (0.31) |
| natalizumab |  | 82 (12.87) |
| alemtuzumab |  | 2 (0.31) |
| daclizumab |  | 1 (0.16) |
| mycophenolate mofetil |  | 2 (0.31) |
| ocrelizumab |  | 1 (0.16) |
| rituximab |  | 14 (2.2) |
| unknown |  | 112 (17.58) |

Initial analyses compared the distribution of within-person and between-person dissimilarity in overall metabolomic profiles; profiles were largely consistent within individuals; the distribution of dissimilarity between subjects was significantly larger than within-subject dissimilarity, as expected (p<1E-16; **Figure 1**). The distributions of within-person and between-person Mahalanobis dissimilarity were similar when stratified by disease status (e.g. MS vs. HC).

### A. Metabolomics differences between MS and HC

#### A.1 Metabolic dysfunction score in overall metabolomic profiles

Overall, we detected shifts in age- and sex-adjusted metabolomic profiles in people with MS relative to HC (Figure 2; p=4.89E-7) using our age- and sex-adjusted metabolomic dysfunction score. We identified samples which were highly divergent from the reference population (defined as >90^th^ percentile of the metabolomic dysfunction score in HC); 131 (20.56%) MS patients had metabolomic dysfunction scores greater than this threshold, providing additional evidence of divergence in overall metabolite profiles (p=7.65E-5). Results were consistent when we included only patients who were not on a disease modifying therapy (DMT) at the time of blood collection, suggesting that the observed differences were not driven by DMT exposure. None of the metabolites differed across therapy classes after adjusting for false discovery (**Supplemental Table 2; Supplemental Figure 3**).

**Figure 2.**
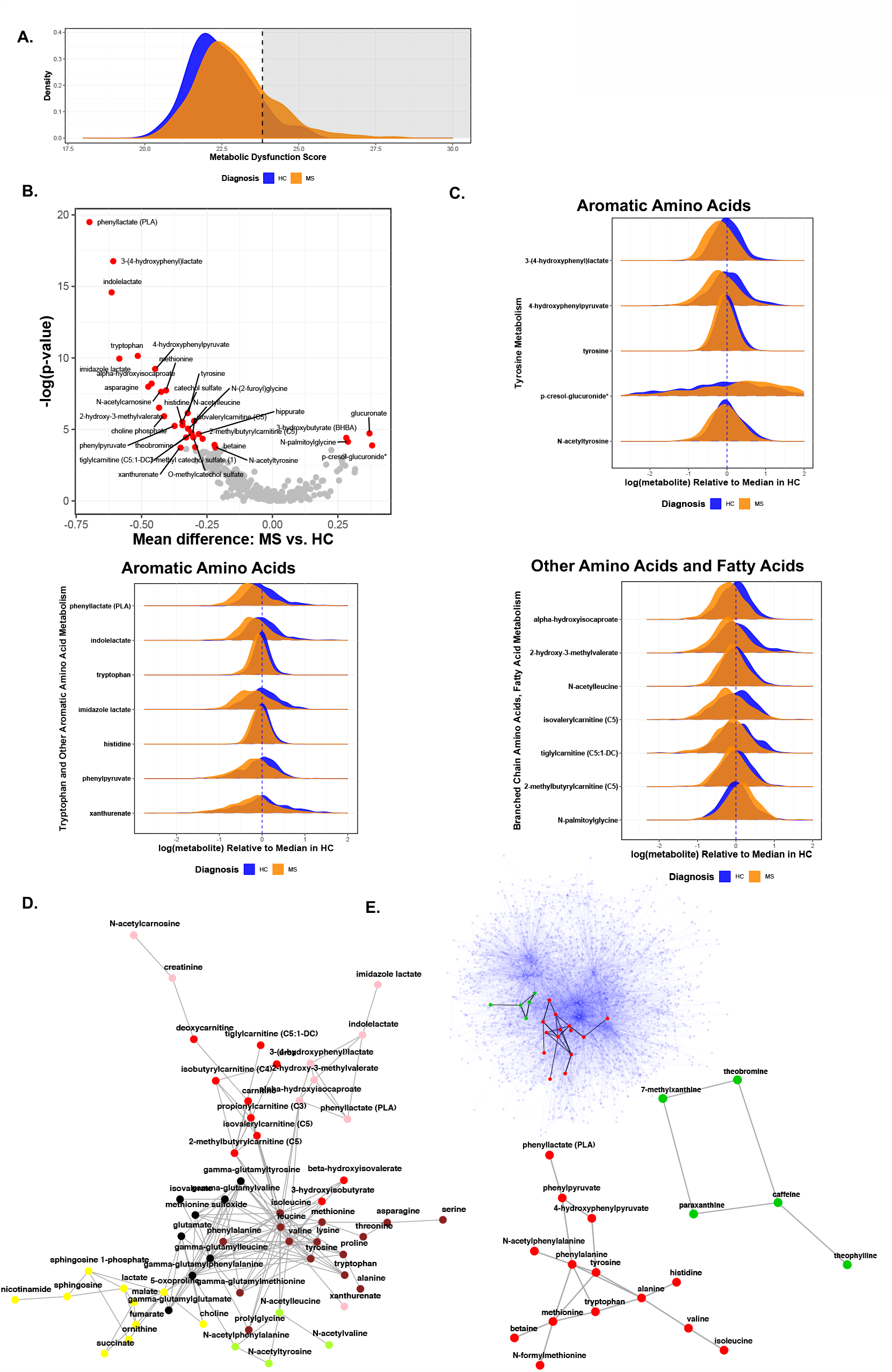
Differences in metabolomic profiles between MS and HC. **A**. Differences in overall metabolomic profiles. The dotted line denotes the 90^th^ percentile of metabolomic dysfunction in HC. **B**. Volcano plot of results of individual metabolites. Each point denotes one metabolite. The x-axis denotes the mean difference in metabolite levels between MS patients and HC, while the y-axis denotes the - log(p-value) for a test of the difference between MS and HC. Red-colored and labeled metabolites denote those significantly different between MS and HC (FDR-adjusted p-value < 0.05). **C**. The distribution of metabolites that significantly differed between MS vs. HC (FDR-adjusted p-value < 0.05); plotted distributions are scaled to the median within HC. **D**. A largest connected component of the agnostic metabolite network derived using WGCNA. Each node represents a metabolite; the color of the node represents the module color, as labeled by WGCNA. The brown, green, yellow, pink and red modules are significantly different between MS patients and HC. **E**. The blue network represents all Metacyc metabolic reactions; nodes represent a metabolite and edges represent a metabolic reaction (e.g. reactant-product connection). We mapped results of the individual metabolites which differed between MS vs. HC with FDR <0.25 and extracted subnetworks of enriched metabolite interactions (corresponding roughly to metabolic pathways) using Prize-collecting Steiner Forest (PCSF) graph optimization. The bottom networks represent the two largest subnetworks extracted after PCSF optimization.

**Table 2.** Analyses of metabolomic differences between multiple sclerosis (MS) and healthy controls (HC) using WGCNA.

| Module* | Overview of Module Composition | Module Size** | Including all samples (n=954) |  |  | Restricting to RRMS patients on no therapy and HC (n=434) |  |  |
| --- | --- | --- | --- | --- | --- | --- | --- | --- |
|  |  |  | Mean difference between MS vs. HC (95% CI)*** | P value | FDR-adjusted P value | Mean difference between MS vs. HC (95% CI)*** | P value | FDR-adjusted P value |
| pink | Aromatic Amino Acids; Branched Chain Amino Acids | 12 | -2.12 (-2.58, -1.65) | 2.13E-19 | 2.77E-18 | -1.87 (-2.51, -1.22) | 1.55E-08 | 2.02E-07 |
| brown | Other Amino Acids | 22 | -1.16 (-1.64, -0.69) | 1.29E-06 | 1.55E-05 | -1.43 (-2.09, -0.78) | 1.65E-05 | 1.99E-04 |
| turquoise | Xanthine metabolites | 26 | -0.97 (-1.43, -0.50) | 4.17E-05 | 4.59E-04 | -0.96 (-1.67, -0.24) | 8.75E-03 | 8.75E-02 |
| green yellow | Acetylated Amino Acid Derivatives | 7 | -0.96 (-1.41, -0.50) | 4.24E-05 | 4.59E-04 | -0.81 (-1.53, -0.09) | 2.65E-02 | 2.39E-01 |
| red | Branched chain amino acids; carnitine metabolites | 14 | -0.93 (-1.39, -0.47) | 7.94E-05 | 7.15E-04 | -0.96 (-1.63, -0.28) | 5.55E-03 | 6.10E-02 |
| magenta | Bile Acid Metabolites | 9 | -0.78 (-1.25, -0.32) | 9.36E-04 | 7.48E-03 | -0.75 (-1.45, -0.05) | 3.52E-02 | 2.81E-01 |
| blue | Fatty acid metabolites; Acyl Carnitine metabolites | 25 | 0.52 (0.05, 0.99) | 2.97E-02 | 2.08E-01 | 0.68 (-0.03, 1.39) | 5.87E-02 | 4.11E-01 |
| tan | Vitamin metabolites | 5 | -0.31 (-0.75, 0.14) | 1.81E-01 | 9.03E-01 | -0.00 (-0.63, 0.62) | 9.94E-01 | 1.00E+00 |
| yellow | Steroid metabolites | 21 | 0.12 (-0.34, 0.58) | 6.11E-01 | 1.00E+00 | 0.43 (-0.23, 1.09) | 2.01E-01 | 1.00E+00 |
| purple | Aromatic Amino Acids; Benzoate Metabolism | 7 | -0.07 (-0.53, 0.38) | 7.48E-01 | 1.00E+00 | 0.07 (-0.62, 0.75) | 8.46E-01 | 1.00E+00 |
| black | Gamma glutamyl metabolites | 12 | 0.06 (-0.38, 0.51) | 7.80E-01 | 1.00E+00 | -0.28 (-1.00, 0.44) | 4.48E-01 | 1.00E+00 |
| green | Methionine, Glycine Metabolism-related metabolites | 16 | -0.05 (-0.51, 0.41) | 8.30E-01 | 1.00E+00 | -0.18 (-0.82, 0.45) | 5.76E-01 | 1.00E+00 |

\*Module name denotes arbitrary name of set of metabolites identified by WGCNA.
\*\*Module size denotes the number of metabolites in a given module.
\*\*\*Mean differences are adjusted for age, sex and race; values displayed denote mean differences eigenmetabolite levels scaled by a factor of $10^2$

#### A.2. Differences in individual metabolites and metabolic pathways

In analyses using WGCNA, we found strong reductions among MS patients in lactate-related metabolites in aromatic amino acid (AAA) pathways (e.g. tryptophan, phenylalanine metabolism). For example, phenyllactate (PLA), 3-(4-hydroxyphenyl)-lactate, indolelactate, and imidazole lactate were significantly reduced in people with MS (all FDR-adjusted p<1E-8; **Supplemental Table 3**). Similarly, in analyses where we grouped metabolites a priori based on related biologic functions (e.g. glutathione metabolism, tryptophan metabolism, among others), we identified highly statistically significant differences in metabolic pathways including other AAA metabolites (**Figure 2; Table 2;** all FDR-adjusted p <1E-4). These results were consistent across different types of pathway and network analyses (**Supplemental Table 4**). We also found strong reductions in branched-chain amino acid (BCAA)-related metabolites in both individual and pathway-based analyses. We observed differences in metabolite pathways related to bile acid metabolism (FDR-adjusted p=3.14E-3), xanthine metabolites (FDR-adjusted p=4.91E-5) and acetylated amino acids (FDR-adjusted p=4.19E-04). In analyses using compounds and reactions from the MetaCyc(30) reaction database (as a proxy for estimated enzyme activity), we also identified a network of amino acids (predominantly AAA) and xanthine (caffeine) metabolites enriched in people with MS. Likewise, we also noted alterations in ratios of tyrosine (reactant) to 4-hydroxyphenylpyruvate (product) and phenylalanine to phenylpyruvate in people with MS, suggesting reduced activity of enzymes involved in these metabolic reactions (**Supplemental Table 5**). Similar to metabolic dysfunction analyses, results were relatively consistent when we included only patients who were not on a disease modifying therapy (DMT) at the time of blood collection.

**Table 3.**
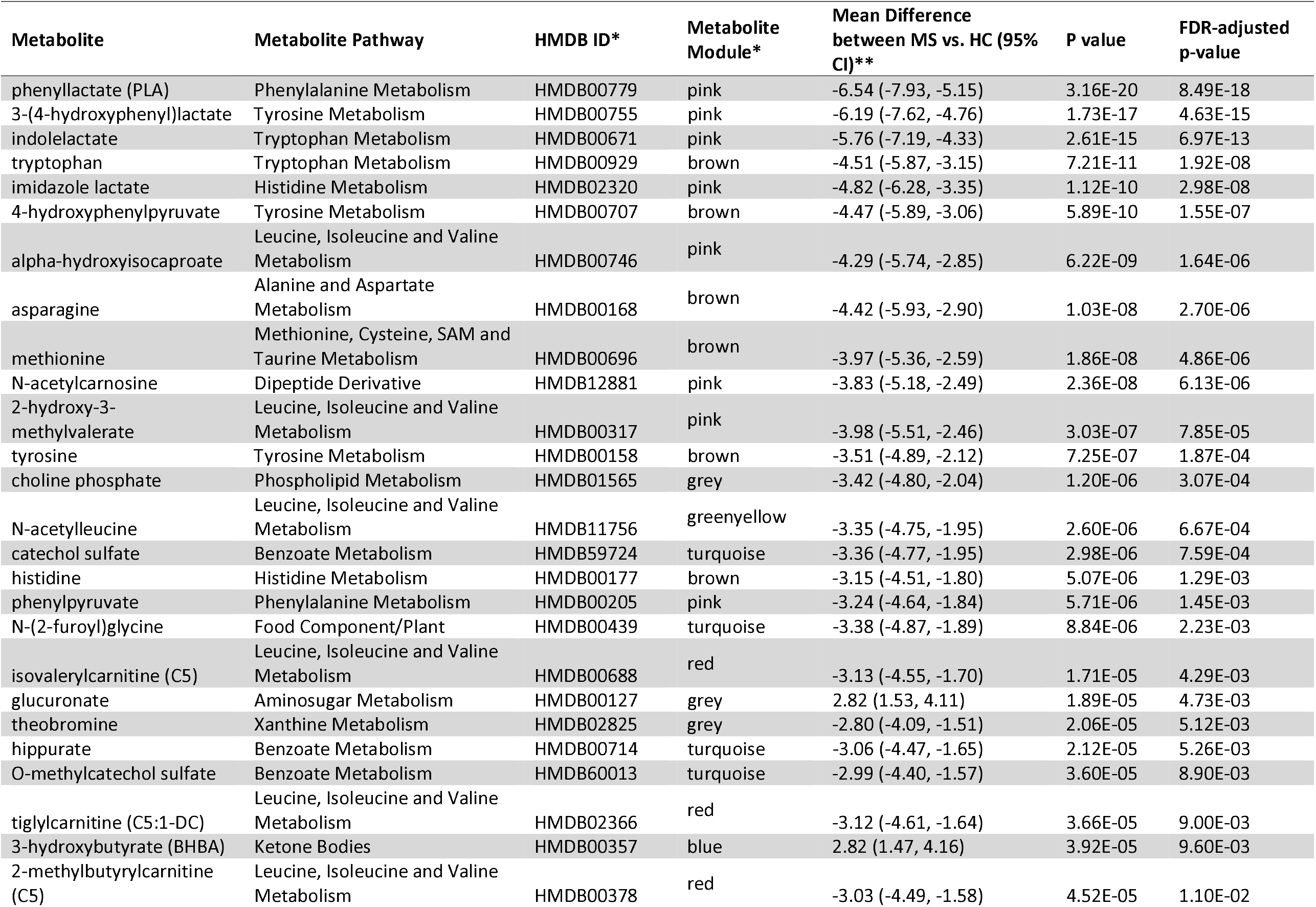
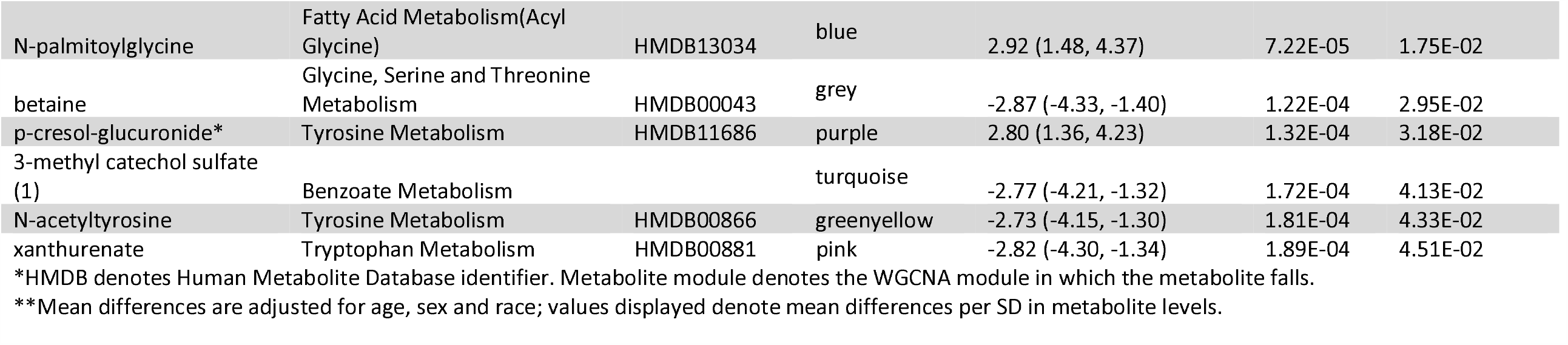
Analyses of metabolomic differences between multiple sclerosis (MS) and healthy controls (HC) for individual metabolites with FDR-adjusted p-values <0.05.

### B. Association of metabolomics profiles with MS characteristics and disease severity measures

Higher levels of metabolic dysfunction were associated with increased disability status; MS patients using a cane had the highest dysfunction scores (**Figure 3**). In analyses assessing the association between individual metabolites and EDSS scores, we observed that reductions in AAA-related metabolites were associated with higher EDSS scores independent of age (**Figure 3; Table 3; Supplemental Table 6**). Specifically, reduced levels of 3-(4-hydroxyphenyl)lactate (tyrosine metabolism) were significantly associated with higher EDSS scores (FDR-adjusted p-value <0.05). Reductions in other AAA metabolites (indolelactate, imidazole lactate, phenyllactate [PLA], kynurenine, kynurenate, tryptophan, and phenylalanine) were also nominally associated with higher EDSS scores (unadjusted p-value <0.05; **Supplemental Table 7**). Several of these metabolites are largely produced by reductive pathway of gut microbial metabolism of AAA. In network or pathway-based analyses, we also observed consistent significant associations between AAA pathways and EDSS scores (**Supplemental Table 8**). Additionally, increases in multiple gut-microbiota derived AAA metabolites (which are also uremic metabotoxins) — p-cresol glucuronide, p-cresol sulfate and phenylacetylglutamine — were associated with greater disability (**Figure 3, Supplemental Table 7**). These metabolites are derived from the oxidative pathway of gut microbial metabolism of AAA.(33) To assess whether the balance between the reductive and oxidative metabolism of AAA in the gut was associated with MS and disease severity, we utilized ratios of metabolites derived from these pathways for individual amino acids (**Figure 3E**). We noted highly significant associations between the ratio of oxidative to reductive metabolites of phenylalanine (phenylacetylglutamine to phenyllactate ratio), tyrosine (p-cresol glucuronide or sulfate to 3-(4-hydroxyphenyl)lactate) and tryptophan (indole acetate to indolelactate ratio) and EDSS scores, suggesting a shift in reductive versus oxidative metabolism of AAA may be related to disability (**Figure 3F**).

**Figure 3.**
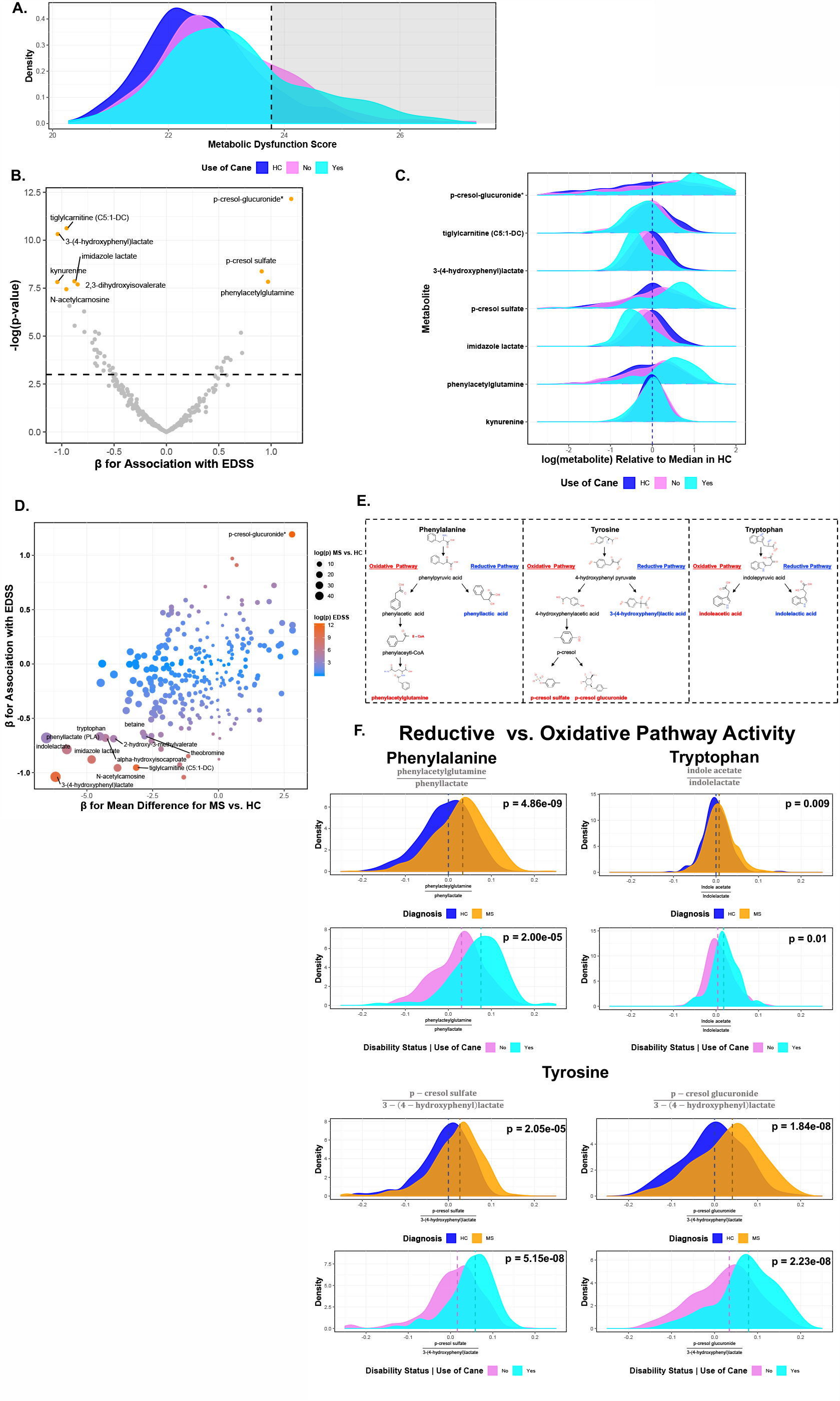
Association between metabolomic profiles and disability in people with MS. **A**. Differences in overall metabolomic profiles by disability status (use of cane vs. no cane). The dotted line denotes the 90^th^ percentile of metabolomic dysfunction in HC. **B**. Volcano plot of results of individual metabolites; the x-axis denotes the association between metabolite level and EDSS scores, while the y-axis denotes the - log(p-value) for a test of association between metabolite level and EDSS score. The dotted horizontal line denotes nominal significance (e.g. -log[p-value=0.05].). **C**. The distribution of metabolites that were potentially associated with EDSS scores by disability status (FDR-adjusted p-value < 0.15); plotted distributions are scaled to the median within HC. **D**. The relationship between metabolite differences between MS vs. HC and association between metabolite level and disability; each point represents a metabolite. The size of point denotes the -log(p-value) for a test of the difference between MS and HC while the color denotes the -log(p-value) for how strongly the metabolite is associated with EDSS. Darker red colors denote stronger levels of association with EDSS. Metabolites which are labeled are metabolites that are significantly different between MS and HC (FDR-adjusted p-value < 0.05) and are at least nominally associated with EDSS score (unadjusted p-value < 0.05). **E**. Oxidative and reductive pathway metabolism of aromatic amino acids, adapted from Dodd et al.(33) Metabolites colored in blue denote reductive pathways metabolites (and are the denominators for ratios plotted included in **Figure 3 F**). Metabolites colored in red denote oxidative pathway metabolites (and are the numerators for ratios plotted in **Figure 3 F**). **F**. The association between oxidative vs. reductive pathway metabolism for aromatic amino acids. For each test, we modeled the ratio of oxidative terminal metabolites to reductive terminal metabolites, as identified from previous studies. The top plot for each ratio denotes differences between MS patients and HC, while the bottom panel denotes the difference in the ratio of metabolites between MS patients using a cane (e.g. having a high level of disability) relative to those not using a cane (e.g. lower levels of disability). P-values are derived from GEE models adjusting for age, sex and race. For models of disability status, we evaluated the association between metabolite ratios and continuous EDSS using a similarly adjusted GEE model.

These results were also relatively consistent in the subset of participants with OCT imaging. AAA metabolites (xanthurenate, p-cresol-glucuronide*, 4-hydroxyphenylpyruvate, phenylacetylglutamine, and phenyllactate) and metabolite ratios (phenylacteylglutamine to phenyllactate and p-cresol glucuronide to 3-(4-hydroxyphenyl)lactate) were also associated with differences in GCIPL thickness (**Supplemental Table 9**). Finally, results were also consistent when we used the ARMSS (rather than EDSS) as a measure of disease-severity.

We also noted a significant positive association between the magnitude of the difference between MS and HC for a given metabolite and the magnitude of the association between individual metabolite level and EDSS scores (correlation [β_MS vs. HC_,β_EDSS_] = 0.41; p=1.35E-12). For example, the metabolite p-cresol-glucuronide is elevated in people with MS relative to HC; higher levels are also associated with higher levels of disability (**Figure 3**).

### C. Sensitivity analyses for primary metabolomics analyses between MS vs. HC and disease severity measures

The adjusted mean difference between MS and HC in a given metabolite level was not associated with how strongly a metabolite correlated with age in HC (correlation [β_MS vs. HC_, β_Age in HC_]= −0.09; p=0.16; **Supplemental Figure 4**). We also performed a set of sensitivity analyses further evaluating the robustness of our results. Results were relatively consistent when we adjusted for BMI when available and for current DMT use (for analyses of disability), and after restricting analyses to individuals not currently taking DMTs. Results were also consistent when repeating analyses using a leave-one-out procedure excluding individual batches and repeated all analyses in the remaining set. Results were also similar in stratified analyses by serum versus plasma.

### D. Single cell metabolic gene expression in MS vs. HC in blood and CSF

Because of the strong differences in amino acid metabolism (specifically in AAA metabolism) identified using circulating metabolomics, we concentrated our snRNA-seq analyses of pathway differences in cell-type specific gene expression between MS and HC in blood and CSF on these metabolic pathways, in particular (**Figure 4**). We identified significant differences in pathway activity of AAA metabolic pathways between MS and HC in monocyte cell clusters, including both clusters of cells enriched in blood and CSF; people with MS tended to have significantly lower levels of pathway activity in theses cell types.

**Figure 4.**
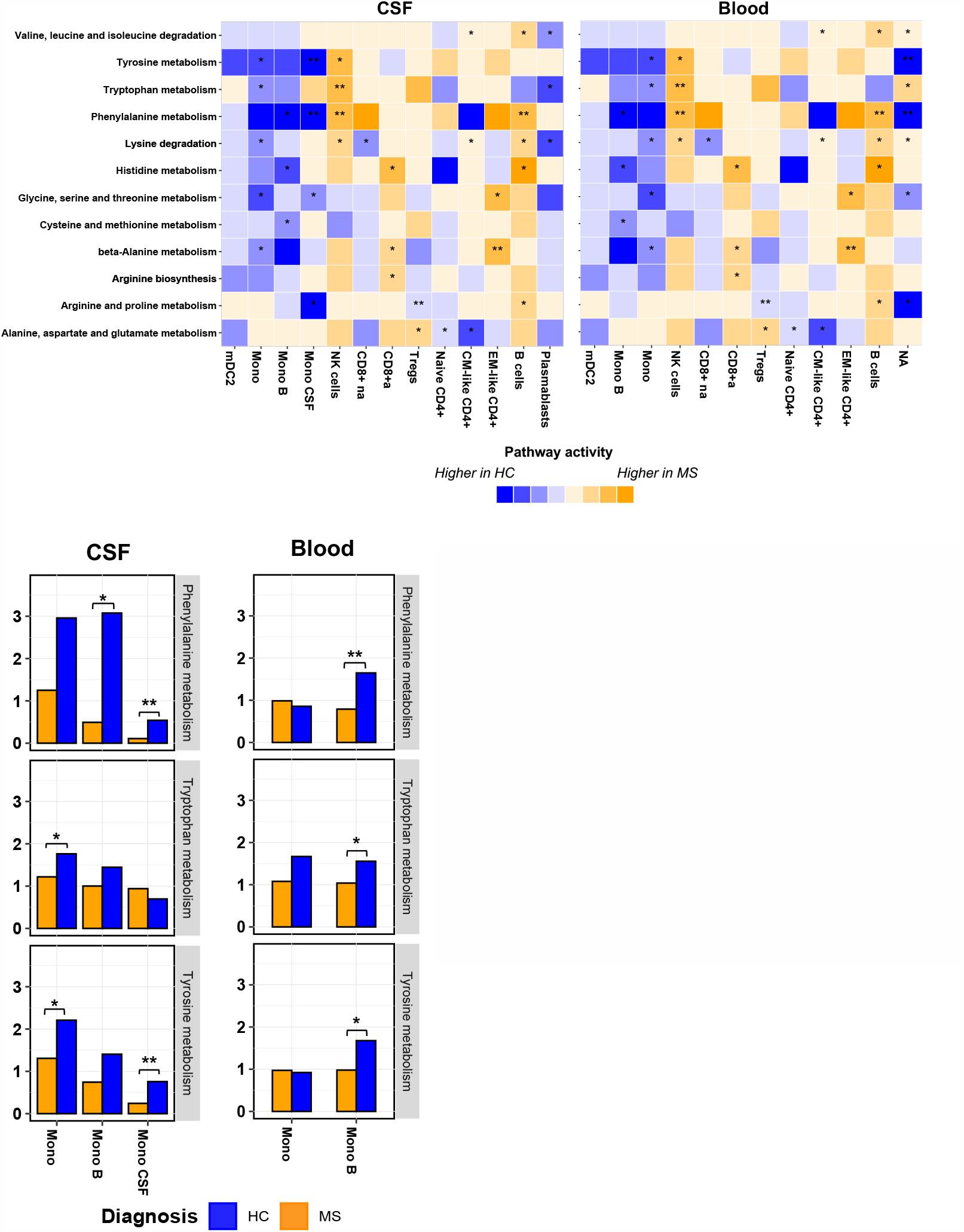
Differences in pathway activity scores between MS vs. HC by cell type. **A**. Heatmap of amino acid pathway activity differences between MS and HC across cell types. The color of the cell denotes the degree of difference in pathway activity scores; darker blue cells denote pathway activity scores which are higher in HC relative to MS whereas darker orange cells denote pathway scores that are higher in MS relative to HC. A single star (*) denotes p-value <0.05, (**) denotes p-values <0.01. **B**. Differences in pathway activity scores for monocyte-like cell clusters for aromatic amino acid pathways between MS and HC samples from CSF (left) and blood (right). As above, a single star (*) denotes p-value <0.05, (**) denotes p-values <0.01.

### E. Effects of AAA-derived metabotoxins on human monocytes

We tested the effects of AAA-derived metabotoxins – IAA, PAG, PCS and PCG, that were identified as being related to higher EDSS severity and lower GCIPL thickness, on human peripheral blood mononuclear cells from healthy controls. We noted an increase in TNFα production from CD14^high^ monocytes with IAA treatment with a clear dose response relationship (**Figure 5**). In a subset of participants, we also noted an increase in IL-6 production from CD14^high^ monocytes with either PAG or IAA treatment (**Supplemental Figure 5**).

**Figure 5.**
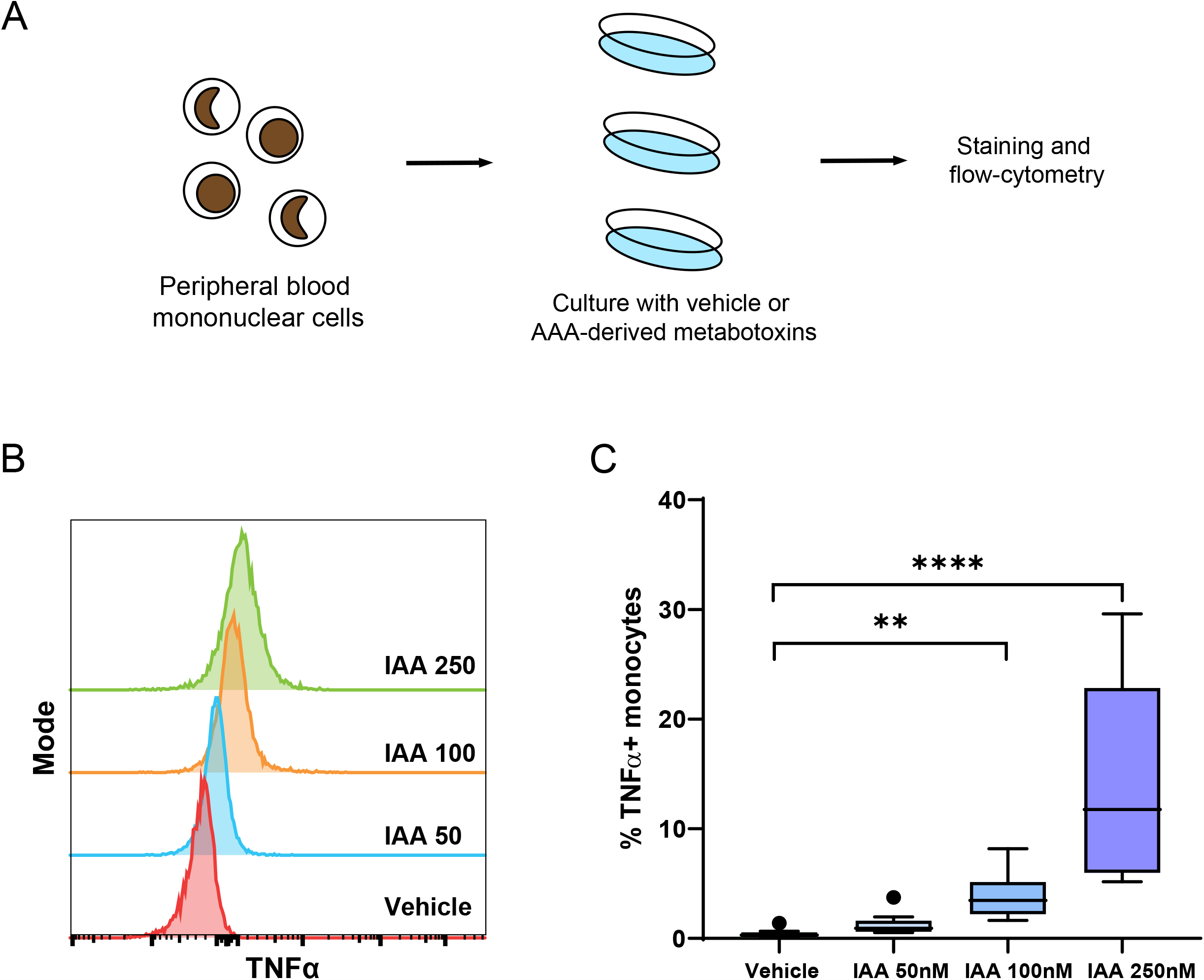
Effects of AAA metabotoxins on human monocytes. A. We treated freshly isolated peripheral blood mononuclear cells from healthy controls with either vehicle or escalating doses of various metabotoxins identified from our initial metabolomics analyses for 24 hours. B. We then utilized multiparametric flowcytometry to evaluate the change in pro-inflammatory cytokine production from monocytes (CD14^high^) and noted an increase in TNFα production with increasing doses of IAA. C. Quantification of increase in the proportion of TNFα producing monocytes with IAA treatment.

## DISCUSSION

This large-scale study identified differences in metabolomic profiles in people with MS relative to HC using a set of individual and pathway-based analyses. Namely, several AAA were altered in MS and lower levels of AAA metabolites derived from the reductive metabolic pathway, and increased levels of AAA metabolites derived from the oxidative pathway, were associated with higher disability scores (**Supplemental Figure 6**). Additionally, analysis of single-cell RNA sequencing data from blood and CSF derived immune cells demonstrated altered AAA metabolism in MS compared to controls. Finally, the identified AAA metabolites linked to higher disability had functional effects on human monocytes and increased production of TNFα in this population of immune cells.

We identified a marked disruption of multiple amino acid metabolic pathways – especially in downstream metabolism of several AAA that are primarily derived from gut microbial reactions (**Figure 3**). Specifically, we noted a broad shift in AAA towards oxidative pathway metabolites relative to reductive pathway metabolites (**e**.**g. Figure 3E**). For example, reductive pathway metabolites (e.g. 3-[4-hydroxy]phenylactate [tyrosine], indole lactate [tryptophan], phenyllactate [phenylalanine]) were largely reduced in people with MS. These specific AAA metabolites are found in large quantities in fermented foods and were recently identified as agonists for the HCA_3_ receptor, which is highly expressed on innate immune cells in humans and is thought to be involved in anti-inflammatory responses (34). Furthermore, these metabolites (e.g. indolelactate) can serve as agonists for the AhR and mediate immunosuppressive actions in immune and glial cells (e.g. limiting pathogenic activities of astrocytes), lower levels could result in increased inflammatory disease activity, both in the peripheral immune system and within the CNS. Consistent with observations that an imbalance between these two pathways has a role in MS, we noted strong associations between oxidative AAA pathway metabolites and increased MS risk and disease severity. Intriguingly, these results were consistent in analyses of metabolic pathway-level changes gene expression in several cell types – most notably in a cluster of CSF-enriched monocytes whose marker gene signature resembled homeostatic microglia.(24, 35).

Phenylacetylglutamine, derived from phenylacetate, is a product of the gut microbial oxidative metabolism of phenylalanine (33, 36). A recent study demonstrated that phenylacetylglutamine (phenylalanine oxidative product) is associated with the risk of cardiovascular disease due to a direct effect on platelet reactivity through adrenergic receptor signaling (36). Similarly, the products of oxidative metabolism of tyrosine (p-cresol sulfate and p-cresol glucuronide) and tryptophan (indole acetate) are also metabotoxins associated with inflammation promotion (37). They are altered in other neurologic diseases and can increase the risk for cardiovascular disease and cardiac dysfunction (36). Several epidemiologic studies in MS have demonstrated a negative impact of vascular and related comorbidities on MS disease severity, and these metabolic alterations may serve as a link between the two conditions (38). Intriguingly, specific bacterial genes are required for reductive vs. oxidative (*por*A for the oxidative pathway and *fldH* for the reductive pathway); future studies will explicitly link shifts in AAA with changes in theses candidate metagenomic genes (33). Functional analyses also demonstrated that treatment with indole acetate led to increased production of the pro-inflammatory cytokine TNFα from human monocytes. TNFα has an important role in MS disease pathogenesis(39) and the effect of this metabolite on TNFα production from myeloid cells provides a possible mechanism by which alterations in the metabolome could lead to worsened MS disease activity and severity.

Also consistent with prior studies, we noted changes in other tryptophan pathway metabolites, with lower circulating levels of both circulating tryptophan and its endogenous- (e.g. kynurenine)-derived metabolites in MS compared to HC (9, 40). As described above, some tryptophan metabolites limit CNS inflammation via AhR-mediated mechanisms in both microglia and astrocytes in the experimental autoimmune encephalitis (EAE) model of MS; microglial AhR deletion worsened EAE, increasing demyelination and CNS monocyte recruitment. Levels of related amino acid metabolites were also reduced in people with MS; the source of these changes remains unclear: diet, altered gut microbiota metabolism of these amino acids, or increased consumption by activated immune cells may all contribute. For example, mechanistic studies have shown that in mice lacking the PD-1 inhibitory checkpoint receptor on T-cells, systemic decreases in tryptophan and tyrosine were due to increased uptake by activated T-cells (41). Downstream reductions in circulating levels of these specific amino acids led to a deficiency in the neurotransmitters serotonin and dopamine in the brain and resulted in increased anxiety-like behaviors. Mood disorders like depression and anxiety are very common in people with MS; it is possible that reduced levels of these metabolites could be a contributing cause of these common comorbidities in MS (42).

Several other metabolic pathways including BCAA, bile acid metabolism and xanthine metabolism were altered in MS compared to controls and have previously reported in a smaller study of people with MS or in animal models. Circulating levels of BCAAs were reduced in MS. BCAAs are critical for the maintenance of regulatory T cells, and their suppressive function *in vivo*. Hence, lower BCAA levels could potentially impair Treg number and function and predispose to increased inflammatory T cell activity (43). We recently linked observations of reduced bile acids in people with MS to key underlying mechanisms and found that bile acid supplementation prevented the polarization of astrocytes and microglia to neurotoxic phenotypes in EAE models (44). Here, we also noted changes in xenobiotic metabolism – including xanthine/caffeine metabolism in MS. Caffeine metabolism is altered in other neurodegenerative disorders, and intake of caffeine has been linked to the risk of developing MS as well as disease severity; our results may suggest that altered caffeine metabolism may underlie the observed associations. In complementary analyses, mapping identified metabolites to a network created from known metabolic reactions, we identified two networks of metabolites that were enriched in MS compared to controls – these again included a network of multiple AAA and BCAAs and a network of caffeine metabolites. Thus, both agnostic and pathway-based analyses yielded complementary results.

There are several noteworthy strengths of this study, which include its large sample size, multiple sites, detailed analyses, and comprehensive assessment of circulating metabolites that were performed using an identical metabolomics platform. We also implemented a stringent QC protocol evaluating metabolite stability within- and between-persons and over time. We complemented our primary analyses with extensive sensitivity analyses; we repeated analyses using a leave-one-out procedure to ensure results were not driven by a single site or batch. We were also able to integrate data from other ‘omics sources in novel multi-omics analyses to help guide the interpretation of our findings.

However, there are some limitations. First, the cross-sectional design limits certain conclusions; longitudinal studies are needed to evaluate how metabolomic changes can influence MS risk or disability changes over time. We included individuals who are largely prevalent DMT users. Therefore, we could not optimally assess the effects of initiation of different DMTs on the metabolome; several smaller studies have identified changes in circulating metabolites associated with different DMTs.(15) However, we did observe relatively consistent findings when restricting to patients not currently on DMT or after adjusting for DMT type. We also lacked comprehensive BMI information on all participants; however, results were consistent after adjusting for BMI in the subset of individuals where this information was available. We also did not have information on time of last meal for all participants. A detailed assessment of diet or other potentially relevant comorbidities affecting metabolomic profiles was also not available. Many key metabolites differing between MS patients and HC are derived from gut microbial reactions, and we lacked this information. Future studies will link metabolomic profiles with both composition and metagenomic features. While intriguing, analyses that identified shifts in metabolic gene expression within specific cell types cannot identify whether such changes are a contributing cause or a result of the observed differences in the circulating metabolome.

In summary, we have demonstrated strong differences in metabolomic profiles between people with MS and healthy individuals. Namely, we note consistent alterations across each AAA towards increased oxidative relative to reductive pathway activity (implicating a specific subset of gut microbial alterations) as being associated with both MS risk and disability. These changes suggest a shift toward increased production of metabotoxins with a potential reduction in the production of immunomodulatory metabolites. We also identify altered AAA metabolic gene expression in MS monocytic cell populations in the blood and CSF and demonstrate the direct effects of AAA-derived metabotoxins on human monocytes. Ultimately, this novel metabolomics study has implications both for advancing cross-omics methods to understand the disease as well as providing critical new insights into candidate pathologic mechanisms contributing to MS.

## Data Availability

Anonymized data used for this study may be available from the corresponding author on reasonable request, with the proper data sharing agreements in place.

---

**Supplemental Figure 1.**
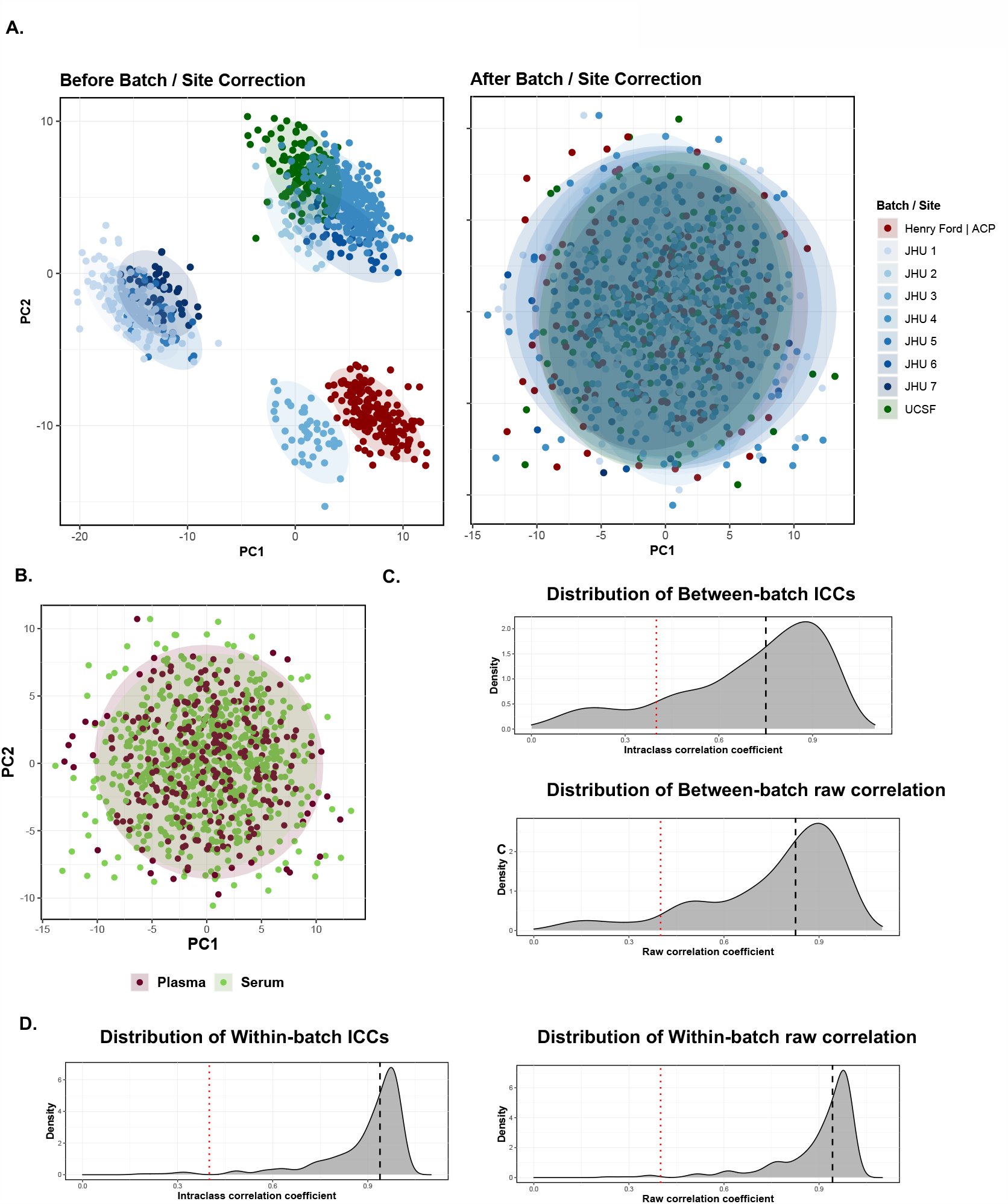
Assessment of combining multiple metabolomics data sets. **A**. PCA plot of metabolite levels before correction for batch (left) and PCA plot after correction for a batch using ComBat. **B**. PCA plot after adjusting for a batch using ComBat by media of metabolomics assessment (plasma or serum). **C**. Distribution of intraclass coefficients (ICC) and raw correlation coefficients for replicate samples between batch samples in which one serum and one plasma sample were included (after application of ComBat). The dotted red line denotes the ICC cut-off for inclusion (>0.40) of a metabolite and the black dashed line denotes the median ICC or raw correlation across all metabolites. The median ICC for serum-plasma replicates is 0.75 (IQR: 0.50, 0.88) and the median raw correlation between serum-plasma replicates is 0.83 (IQR: 0.64, 0.91). **D**. Distribution of ICCs and raw correlation coefficients for duplicate samples included within the same batch (after application of ComBat). The dotted red line denotes the ICC cut-off for inclusion (>0.40) of a metabolite and the black dashed line denotes the median ICC or raw correlation across all metabolites. The median ICC for duplicate samples in the same batch is 0.94 (IQR: 0.86, 0.98) and the median raw correlation between duplicate samples is 0.94 (IQR: 0.88, 0.98).

**Supplemental Figure 2.**
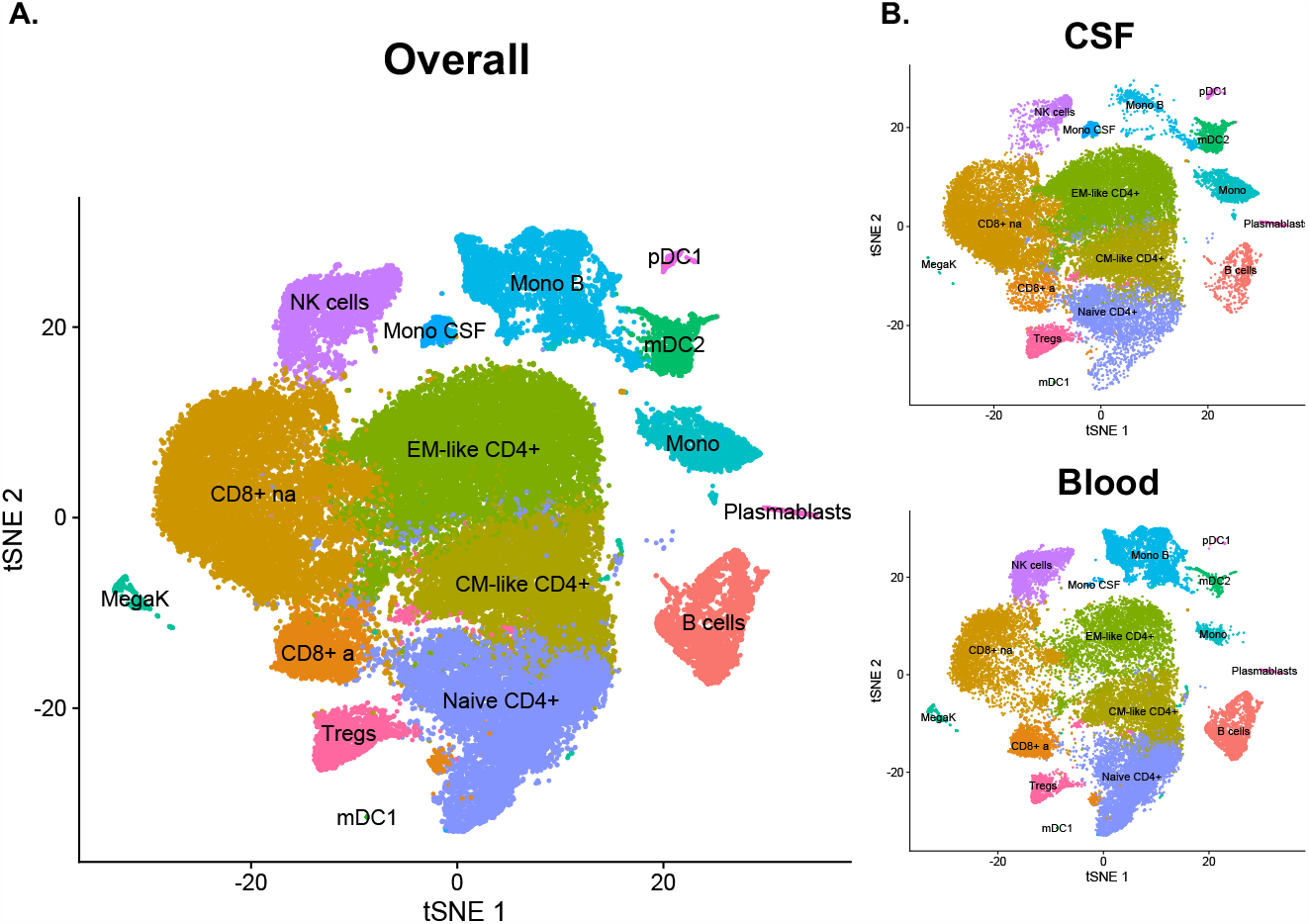
tSNE plots of single cell RNA sequencing data from MS And healthy control blood and CSF. **A**. tSNE plot of cell clustering results including MS and HC and blood and CSF. We identified clusters 15 clusters of different cell types that were identified using marker gene expression. Cell types included Naïve CD4+ T cells (Naïve CD4+), central memory-(CM)-like CD4+ T cells (CM-like CD4+), effector memory-(EM)-like CD4+ T cells (EM-like CD4+), activated CD8+ T cells (CD8+ a), non-activated CD8+ T cells (CD8+ a), T regulatory cells (Tregs), B cells, monocytes (Mono), blood-derived monocytes (Mono B), CSF derived monocytes (Mono CSF), NK cells, myeloid dendritic cells types 1 and 2 (mDC1 and mDC2), plasmacytoid dendritic cells (pDC1), and megakaryocytes (MegaK). **B**. tSNE plot stratified by sample type.

**Supplemental Figure 3.**
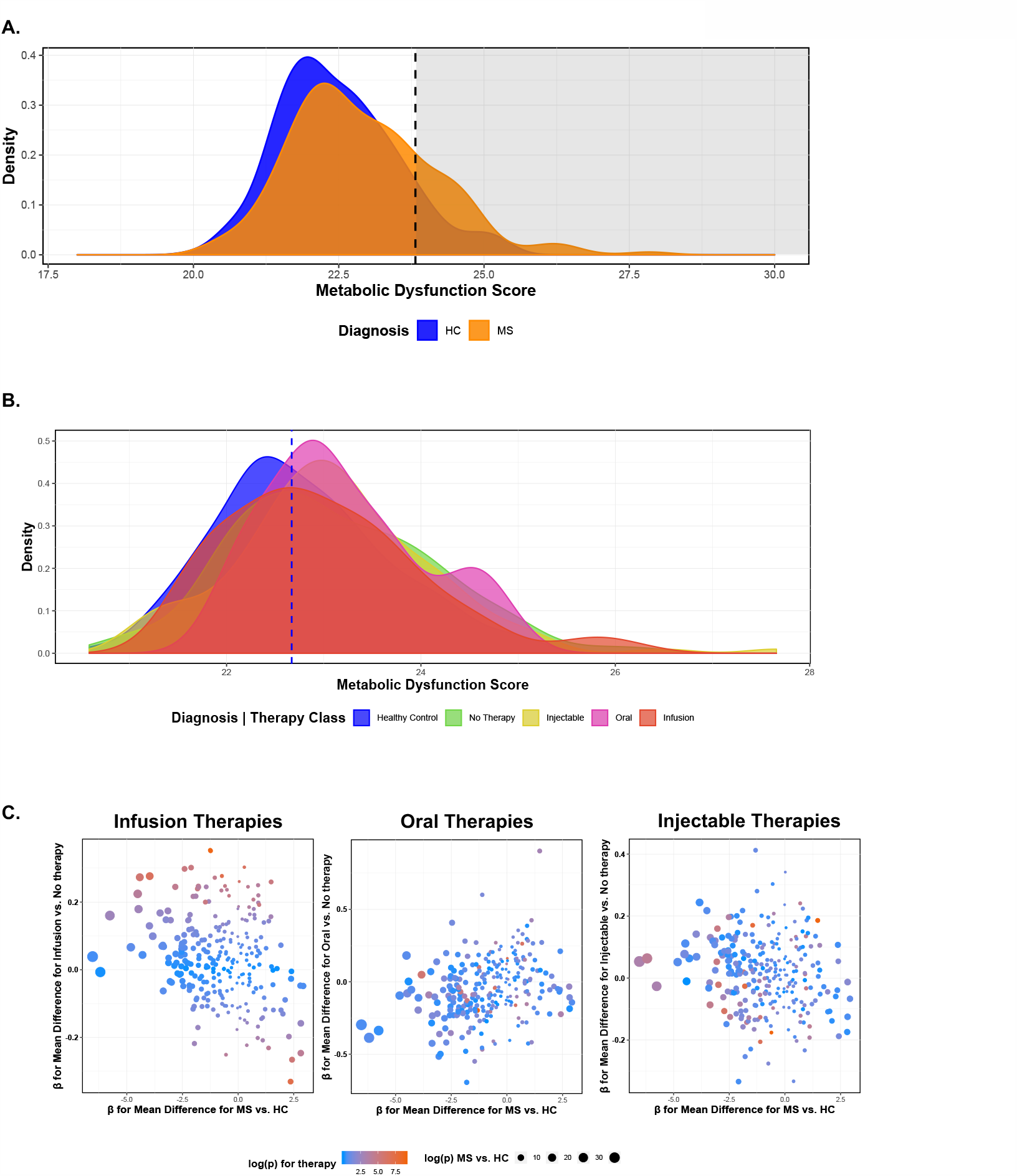
Sensitivity analysis of metabolomics data by treatment status. **A**. Differences in overall metabolomic profiles among untreated MS patients. The dotted line denotes the 90^th^ percentile of metabolomic dysfunction in HC. **B**. Differences in overall metabolomic profiles by MS DMTs. **C**. The relationship between metabolite differences between MS vs. HC and mean differences between individuals who are on infusion, oral or injectable therapies versus those who are not on any therapy. The size of point denotes the -log(p-value) for a test of the difference between MS and HC while the color denotes the -log(p-value) for how strongly the metabolite is associated with each therapy. Darker red colors denote stronger levels of differences between therapy classes. We observed an inverse association between estimated mean differences between MS vs. HC and estimated mean differences between MS patients on infusion or injectable therapies vs. those not on any therapy (for infusion therapies: *r*=-0.26; 95% CI: −0.34, −0.14; p=1E-5; for injectable therapies: −0.16; 95%. CI: −0.27, −0.04; p=0.009). We observed a positive association between estimated mean differences between MS vs. HC and estimated mean differences between MS patients on oral therapies vs. those not on any therapy (*r*=0.29; 95% CI: 0.18, 0.40; p=1E-6). Moderate efficacy/oral therapies included fingolimod, dimethyl fumarate, and teriflunomide. Higher efficacy/infusion therapies included ocrelizumab, rituximab, natalizumab, daclizumab, and alemtuzumab. Lower efficacy/injectable therapies included glatiramer acetate and interferon-beta.

**Supplemental Figure 4.**
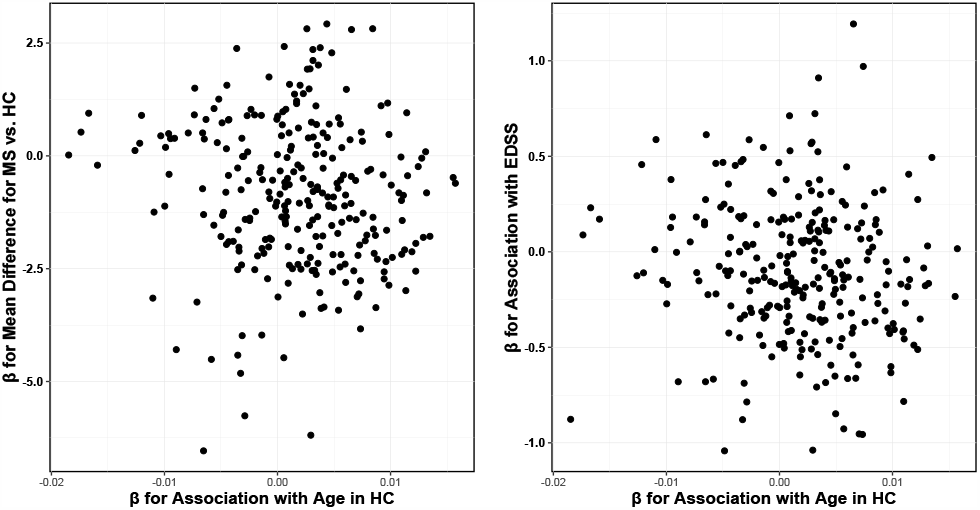
Assessing effect of age on observed relationships between metabolite levels and disease group and severity. The adjusted mean difference in metabolite level between MS and HC (y-axis) versus the magnitude of the association between metabolite level and age (x-axis). We did not observe evidence of an association (correlation= −0.09; p=0.16). The adjusted magnitude of the association between EDSS and metabolite level (y-axis) versus the magnitude of the association between metabolite level and age (x-axis). We did not observe an association (correlation= −0.11; p=0.06)

**Supplemental Figure 5.**
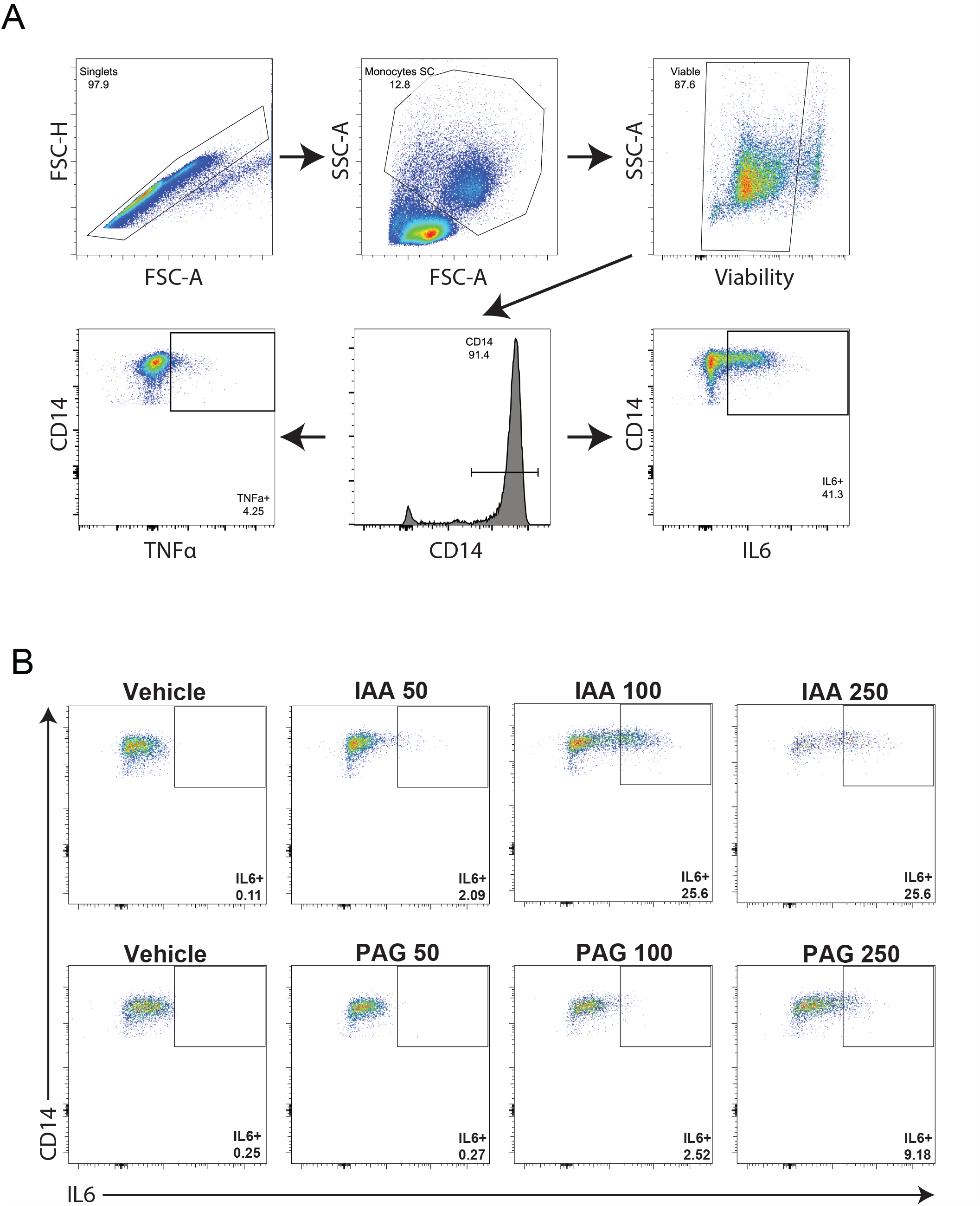
Effect of AAA derived metabotoxins on human monocytes. A. Gating strategy for identifying CD14^high^ monocytes. B. Effect of AAA derived metabotoxins on IL-6 production from human monocytes. A sub-set of participants (n=4) showed increased IL-6 production with IAA and PAG treatment and representative examples are shown in panel B, however no significant effect was noted on IL-6 production with either metabolite in the group as a whole (n=9).

**Supplemental Figure 6.**
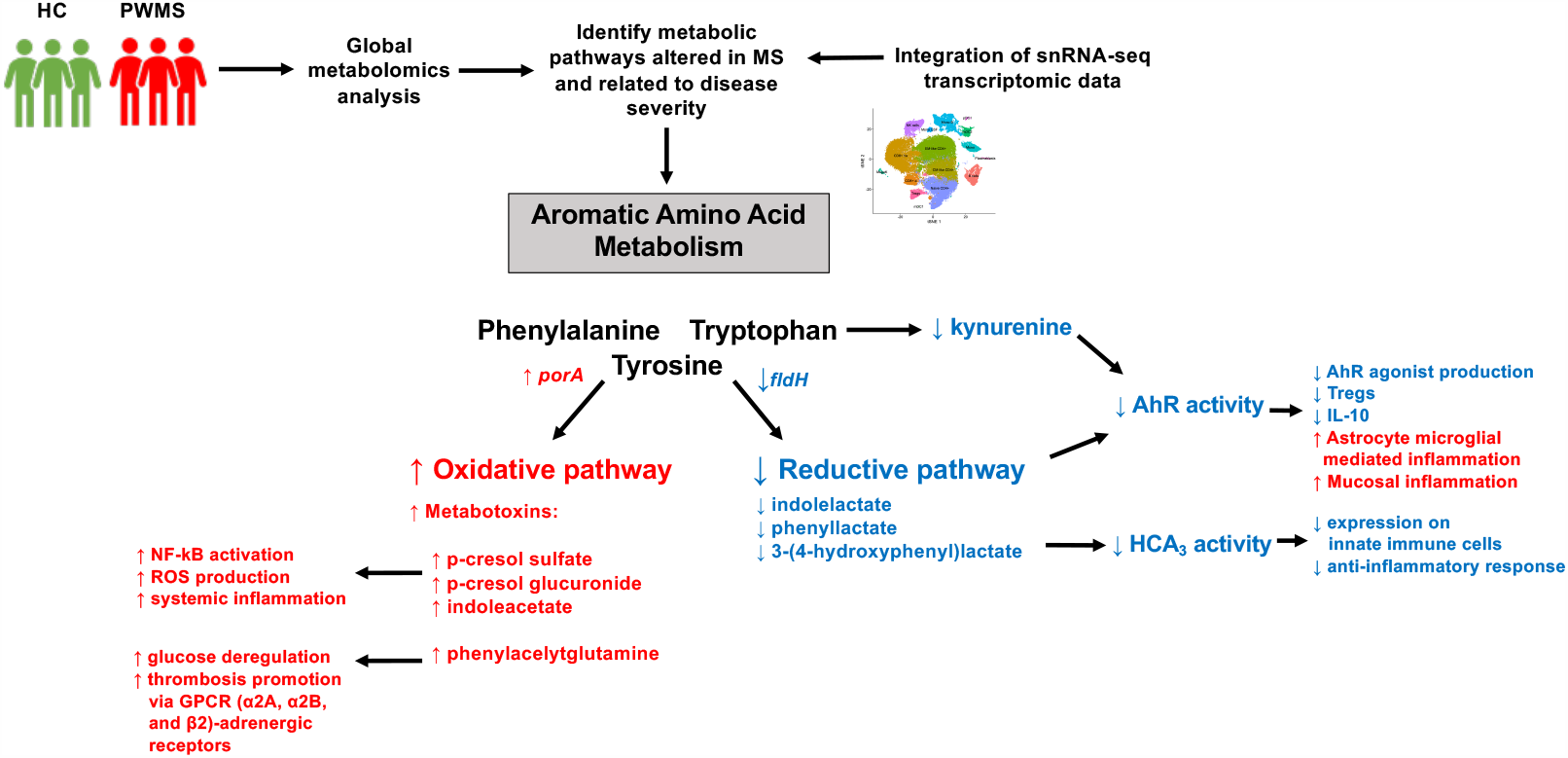
Summary of the results of metabolomics analyses. Our results suggest that disruption in downstream metabolism of several aromatic amino acids (AAA) is associated with MS risk and with disability level in people with MS. We found that reductive pathway metabolites (e.g. 3-[4-hydroxy]phenylactate [tyrosine], indole lactate [tryptophan], phenyllactate [phenylalanine]) were largely reduced in people with MS and shifted towards towards oxidative pathway metabolites (e.g. p-cresol sulfate, p-cresol glucuronide, indoleacetate, and phenylacetylglutamine). Previous evidence demonstrates that lower reductive pathway metabolites are associated with decreases in AhR activity leading to decreases in regulatory T cells, lower IL-10 levels, and increases in astrocyte-mediated microglial inflammation. Lower levels are also associated with reduced HCA3 receptor activity which is highly expressed on innate immune cells in humans and is thought to be involved in anti-inflammatory response. In contrast, higher levels of oxidative pathway metabolites correspond to increases in metabotoxins, which can increase NFkb signaling, reactive oxidative species and systemic inflammation. They also can increase glucose deregulation. Intriguingly, these AAA metabolites are largely the products of gut microbial metabolism, and specific bacterial genes are required for reductive vs. oxidative (porA for the oxidative pathway and fldH for the reductive pathway).

## References

1. Reich DS, Lucchinetti CF, Calabresi PA. Multiple Sclerosis. N. Engl. J. Med. 2018;378(2):169– 180.

2. Ascherio A, Munger KL. Epidemiology of Multiple Sclerosis: From Risk Factors to Prevention-An Update. Semin Neurol 2016;36(2):103–114.

3. International Multiple Sclerosis Genetics Consortium. Multiple sclerosis genomic map implicates peripheral immune cells and microglia in susceptibility. Science 2019;365(6460). doi:10.1126/science.aav7188

4. Patti GJ, Yanes O, Siuzdak G. Innovation: Metabolomics: the apogee of the omics trilogy. Nat. Rev. Mol. Cell Biol. 2012;13(4):263–269.

5. Newgard CB. Metabolomics and Metabolic Diseases: Where Do We Stand?. Cell Metab. 2017;25(1):43–56.

6. Dickens AM et al. A type 2 biomarker separates relapsing-remitting from secondary progressive multiple sclerosis. Neurology 2014;83(17):1492–1499.

7. Bhargava P, Fitzgerald KC, Calabresi PA, Mowry EM. Metabolic alterations in multiple sclerosis and the impact of vitamin D supplementation. JCI Insight 2017;2(19). doi:10.1172/jci.insight.95302

8. Lim CK et al. Kynurenine pathway metabolomics predicts and provides mechanistic insight into multiple sclerosis progression. Sci Rep 2017;7:41473.

9. Nourbakhsh B et al. Altered tryptophan metabolism is associated with pediatric multiple sclerosis risk and course. Ann Clin Transl Neurol 2018;5(10):1211–1221.

10. Shao Y, Le W. Recent advances and perspectives of metabolomics-based investigations in Parkinson’s disease. Mol Neurodegener 2019;14(1):3.

11. Jääskeläinen O et al. Metabolic Profiles Help Discriminate Mild Cognitive Impairment from Dementia Stage in Alzheimer’s Disease. J. Alzheimers Dis. 2020;74(1):277–286.

12. Paley EL. Discovery of Gut Bacteria Specific to Alzheimer’s Associated Diseases is a Clue to Understanding Disease Etiology: Meta-Analysis of Population-Based Data on Human Gut Metagenomics and Metabolomics. J. Alzheimers Dis. 2019;72(1):319–355.

13. Roman Fox S et al. A pilot study evaluating changes in clinical outcomes with weight loss in people with multiple sclerosis 2017;

14. Fitzgerald KC et al. Effect of intermittent vs. daily calorie restriction on changes in weight and patient-reported outcomes in people with multiple sclerosis. Mult Scler Relat Disord 2018;23:33–39.

15. Bhargava P et al. Dimethyl fumarate treatment induces lipid metabolism alterations that are linked to immunological changes. Ann Clin Transl Neurol 2019;6(1):33–45.

16. Bhargava P et al. Multiple sclerosis patients have a diminished serologic response to vitamin D supplementation compared to healthy controls. Mult. Scler. 2016;22(6):753–760.

17. Lang A et al. Retinal layer segmentation of macular OCT images using boundary classification. Biomed Opt Express 2013;4(7):1133–1152.

18. Saidha S et al. Optical coherence tomography reflects brain atrophy in multiple sclerosis: A four-year study. Ann. Neurol. 2015;78(5):801–813.

19. Bhargava P et al. Applying an Open-Source Segmentation Algorithm to Different OCT Devices in Multiple Sclerosis Patients and Healthy Controls: Implications for Clinical Trials. Mult Scler Int 2015;2015:136295.

20. Saidha S et al. Relationships between retinal axonal and neuronal measures and global central nervous system pathology in multiple sclerosis. JAMA Neurol 2013;70(1):34–43.

21. Azary S et al. Contribution of dietary intake to relapse rate in early paediatric multiple sclerosis. J. Neurol. Neurosurg. Psychiatry 2018;89(1):28–33.

22. Pakpoor J et al. Dietary factors and pediatric multiple sclerosis: A case-control study. Mult. Scler. 2018;24(8):1067–1076.

23. Krupp LB et al. International Pediatric Multiple Sclerosis Study Group criteria for pediatric multiple sclerosis and immune-mediated central nervous system demyelinating disorders: revisions to the 2007 definitions. Mult. Scler. 2013;19(10):1261–1267.

24. Schafflick D et al. Integrated single cell analysis of blood and cerebrospinal fluid leukocytes in multiple sclerosis. Nature Communications 2020;11(1):247.

25. Leek JT, Johnson WE, Parker HS, Jaffe AE, Storey JD. The sva package for removing batch effects and other unwanted variation in high-throughput experiments. Bioinformatics 2012;28(6):882–883.

26. Fortin J-P et al. Harmonization of cortical thickness measurements across scanners and sites. Neuroimage 2018;167:104–120.

27. Langfelder P, Horvath S. WGCNA: an R package for weighted correlation network analysis. BMC Bioinformatics 2008;9:559.

28. Lloyd-Price J et al. Multi-omics of the gut microbial ecosystem in inflammatory bowel diseases. Nature 2019;569(7758):655–662.

29. Sato K et al. Visualizing modules of coordinated structural brain atrophy during the course of conversion to Alzheimer’s disease by applying methodology from gene co-expression analysis [Internet]. Neuroimage Clin 2019;24. doi:10.1016/j.nicl.2019.101957

30. Caspi R et al. The MetaCyc database of metabolic pathways and enzymes. Nucleic Acids Res 2018;46(D1):D633–D639.

31. Akhmedov M et al. PCSF: An R-package for network-based interpretation of high-throughput data. PLOS Computational Biology 2017;13(7):e1005694.

32. Xiao Z, Dai Z, Locasale JW. Metabolic landscape of the tumor microenvironment at single cell resolution. Nature Communications 2019;10(1):3763.

33. Dodd D et al. A gut bacterial pathway metabolizes aromatic amino acids into nine circulating metabolites. Nature 2017;551(7682):648–652.

34. Peters A et al. Metabolites of lactic acid bacteria present in fermented foods are highly potent agonists of human hydroxycarboxylic acid receptor 3. PLOS Genetics 2019;15(5):e1008145.

35. Masuda T et al. Spatial and temporal heterogeneity of mouse and human microglia at single-cell resolution. Nature 2019;566(7744):388–392.

36. Nemet I et al. A Cardiovascular Disease-Linked Gut Microbial Metabolite Acts via Adrenergic Receptors. Cell 2020;180(5):862-877.e22.

37. Sankowski B et al. Higher cerebrospinal fluid to plasma ratio of p-cresol sulfate and indoxyl sulfate in patients with Parkinson’s disease. Clin. Chim. Acta 2020;501:165–173.

38. Zhang T et al. Effects of physical comorbidities on disability progression in multiple sclerosis. Neurology 2018;90(5):e419–e427.

39. Fresegna D et al. Re-Examining the Role of TNF in MS Pathogenesis and Therapy. Cells 2020;9(10). doi:10.3390/cells9102290

40. Rothhammer V et al. Type I interferons and microbial metabolites of tryptophan modulate astrocyte activity and central nervous system inflammation via the aryl hydrocarbon receptor. Nat. Med. 2016;22(6):586–597.

41. Miyajima M et al. Metabolic shift induced by systemic activation of T cells in PD-1-deficient mice perturbs brain monoamines and emotional behavior. Nature Immunology 2017;18(12):1342–1352.

42. Marrie RA et al. The incidence and prevalence of psychiatric disorders in multiple sclerosis: A systematic review. Mult Scler 2015;21(3):305–317.

43. Ikeda K et al. Slc3a2 Mediates Branched-Chain Amino-Acid-Dependent Maintenance of Regulatory T Cells. Cell Rep 2017;21(7):1824–1838.

44. Bhargava P et al. Bile acid metabolism is altered in multiple sclerosis and supplementation ameliorates neuroinflammation. J. Clin. Invest. [published online ahead of print: March 17, 2020]; doi:10.1172/JCI129401

